# Study Research Protocol for Phenome India-CSIR Health Cohort Knowledgebase (PI-CHeCK): A Prospective multi-modal follow-up study on a nationwide employee cohort

**DOI:** 10.1101/2024.10.17.24315252

**Authors:** Phenome India Consortium, Shantanu Sengupta

## Abstract

Predicting individual health trajectories based on risk scores can help formulate effective preventive strategies for diseases and their complications. Currently, most risk prediction algorithms rely on epidemiological data from the Caucasian population, which often do not translate well to the Indian population due to ethnic diversity, differing dietary and lifestyle habits, and unique risk profiles. In this multi-center prospective longitudinal study conducted across India, we aim to address these challenges by developing clinically relevant risk prediction scores for cardio-metabolic diseases specifically tailored to the Indian population. India, which accounts for nearly 18% of the global population, also has a significant diaspora worldwide. This program targets longitudinal collection and bio-banking of samples from over 10,000 employees both working and retirees of the Council of Scientific and Industrial Research and their spouses, with baseline sample collection already completed. During the baseline collection, we gathered multi-parametric data including clinical questionnaires, lifestyle and dietary habits, anthropometric parameters, lung function assessments, liver elastography by Fibroscan, electrocardiogram readings, biochemical data, and molecular assays, including but not limited to genomics, plasma proteomics, metabolomics, and fecal microbiome analysis. In addition to exploring associations between these parameters and their cardio-metabolic outcomes, we plan to employ artificial intelligence algorithms to develop predictive models for phenotypic conditions. This study could pave the way for precision medicine tailored to the Indian population, particularly for the middle-income strata, and help refine the normative values for health and disease indicators in India.

## Introduction

Non-communicable diseases (NCDs) contribute significantly to human morbidity and mortality, accounting for 71% of the total 55 million deaths that occur annually worldwide (1). Of these, approximately 17 million deaths occur in individuals younger than 70 years, with 86% of these early fatalities happening in low and middle-income countries (LMICs) (1). India alone accounts for 5.8 million of these deaths, nearly 10% of the global mortality due to NCDs. Notably, the onset of cardiovascular diseases (CVD) in this population occurs nearly a decade earlier (2, 3). Among NCDs, cardiovascular diseases are the leading cause of death, with 17.9 million fatalities, followed by cancer (9.3 million), respiratory disorders (4.1 million), and diabetes (2 million). Conditions like Nonalcoholic fatty liver disease (NAFLD) and nonalcoholic steato-hepatitis (NASH), which also increase cardiovascular risks, are expected to see a rise in global prevalence from 24%- 40% to over 55% by 2040 (4-6).

Given India’s diverse ethnicities and geographic variations, understanding the specific risk factors for NCDs in India is crucial. This is particularly important because India is home to one-sixth of the world’s population and is considered a global genetic melting pot (7). Additionally, a significant portion of the Indian population resides in the diaspora across other countries. Therefore, comprehensive longitudinal studies across India are essential to identify unique risk factors that can guide the development of targeted preventive and personalized health strategies for both the Indian population and its global diaspora. Identifying these factors is crucial for formulating effective policies and guidelines for precision medicine, which enables the prediction of individual health trajectories and allows for early intervention to prevent disease onset or complications. Risk scores have traditionally been used to predict health outcomes based on data from prospective cohort studies. With the advent of multi-omics data and artificial intelligence (AI)-based analytical tools, there are unprecedented opportunities for developing novel personalized risk metrics for predicting health outcomes (8, 9). A longitudinal population study involving baseline phenotypic measurements and long-term follow-up can help elucidate the complex interrelationships between environmental, genetic, and molecular factors and the risk of subsequent diseases. This approach allows for the measurement of parameters well before the disease onset, minimizing selection bias and avoiding issues related to reverse causality. Such epidemiological data are vital for developing risk prediction tools and advancing precision health. Currently, most risk prediction algorithms are based on epidemiological data from Caucasian populations, which are often inaccurate when applied to the Indian context due to ethnic diversity, differing dietary and lifestyle patterns and distinct risk profiles (10-12). Importantly, existing studies on the Indian population are limited in both phenotypic information and diversity.

The Indian population is known to exhibit a predisposition to central obesity, increasing the risk of cardio-metabolic diseases (13, 14). Detailed phenotyping and understanding of these risks’ genetic and metabolic underpinnings is essential. A longitudinal approach that collects phenotypic data across multiple time points (including physiological function, multi-omics data, and microbiome diversity) will enhance understanding of these risks. Integrating various omics data, such as phenome, metabolome, proteome, and genome, is crucial for developing preventive protocols for various NCDs. Phenomics can help discover new risk factors, diagnostic biomarkers and therapeutic targets for precision medicine (15).

To address the risk factors of cardio-metabolic diseases, particularly diabetes and NAFLD, the Council of Scientific and Industrial Research (CSIR) has initiated the multi-center, pan-India study “Phenome India-CSIR Health Cohort Knowledgebase” (PI-CheCK). The study encompasses its 37 constituent laboratories and associated centers across 17 states and 2 union territories (UT) (16) (Figure 1).

**Figure 1:**
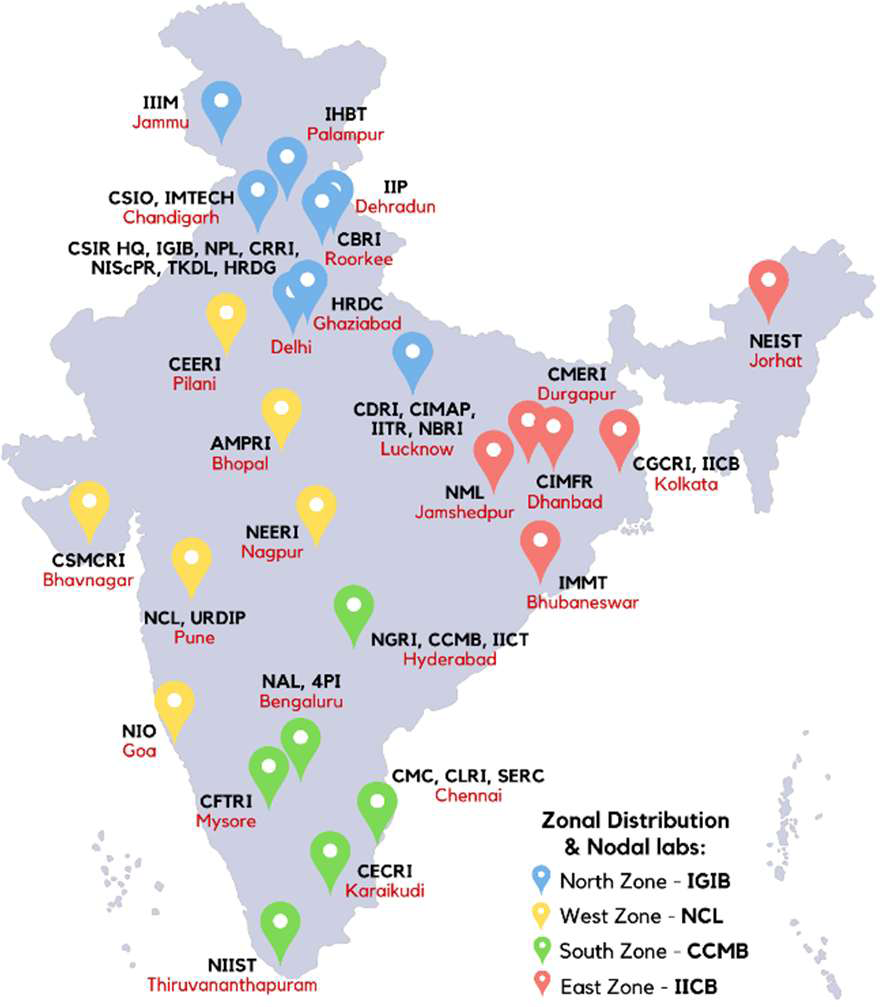
Sampling zones, sites in Phenome India Project, and coordinating labs. Zones are colored and defined for logistics purposes, not in any conventional manner.

CSIR encompasses a diverse range of ethnicities, social-ecological regions and occupational exposures, primarily situated in urban and semi-urban areas which may contribute key factors relevant to this study. At the same time, CSIR staff predominantly come from urban and semi-urban middle- and higher-middle-income groups, resulting in a relatively homogenous socio-economic profile. This planned cohort, comprising approximately 10,000 individuals, includes well-educated working and retired staff as well as their spouses. While India has some notable cohort studies, these are often limited by geographic scope, or the parameters being monitored (6, 17-21). To the best of our knowledge, no nationwide longitudinal cohort of this scale and characterization has been reported before. The study will involve prospective data collection initially over five years at three-time points, gathering extensive health information. This will include but is not limited to:- physiological assessment of lung function via spirometry and oscillometry, cardiac function and heart rate variability (HRV) analysis using echocardiogram (ECG), body composition analysis (BCA), liver function tests through transient elastography (Fibroscan), dermatological assessment of skin moisture, sebum and hydration, as well as anthropometry including muscle strength through dynamometer. In addition, molecular and biochemical assays of blood samples will be conducted, and a comprehensive sample and data repository will be established for multiple omics and microbiome analyses.

The goal is to generate a first-of-its-kind Indian dataset encompassing various health-related and biomedical parameters from a nationwide employee cohort, tracked over several years. This dataset will have significant public health implications, with the potential to identify diagnostic and prognostic biomarkers. An employee cohort is expected to have a lower dropout rate, as participants are educated and literate, which enhances the study’s effectiveness. Moreover, this will promote discussions of clinically relevant findings, ultimately contributing to the development of a healthy and productive scientific workforce at CSIR.

Study Objectives

The primary objectives of the PI-CHeCK study are:

a) To develop a prospective longitudinal cohort of consenting CSIR employees, retirees and their spouses.

b) To develop clinically useful personalized risk prediction scores for cardio-metabolic disorders for pre-diabetes, diabetes, and fatty liver disease.

The long-term objectives for the listed disorders of the study are:

1. To develop risk prediction models for diabetes and liver fibrosis:

(i) Glycemic control: Development of a binary prediction model for the normo-glycemic population to develop:

(a) Diabetes (prior glycated hemoglobin (HbA1c) <5.7 and fasting blood sugar (FBS) <100 to HbA1c ≥6.5 and FBS ≥126 mg/dL (as per American Diabetes Association (ADA) criteria)

(b) Pre-Diabetes (with prior HbA1c <5.7 and FBS <100 to HbA1c between ≥5.7 and <6.5 and FBS ≥100 but less than <126 mg/dL as per ADA criteria)

(ii) NAFLD/Liver Fibrosis: Development of a binary prediction model for F0/F1 (no fibrosis) to F2/F3/F4 (moderate to severe fibrosis/cirrhosis) on liver Fibroscan (Transient Elastography)

2. To create and maintain a database and linked bio-repository/biobank.

## Methods

### Design of the study

The study design is primarily longitudinal with the baseline sample collection phase already being completed and two subsequent follow-up phases planned.

### Sampling sites and sample size determination

All 37 CSIR labs and associated centers were organized into four zones to streamline logistics management. These zones were defined based on the geographical locations of the labs. The baseline number of regular employees at each lab was used as a reference, and this number was multiplied by four to account for the spouses of current employees, an equal number of pensioners, and the spouses of pensioners. The assumptions made were a) the number of pensioners is at least equal to the number of employees in each lab, and b) both employees and pensioners have spouses. This approach provided a total count for each lab, which was then adjusted to ensure that each zone contained at least 2,500 samples. Information on the four zones and their respective labs is provided in Supplementary Table S1. Four zonal nodal Institutes were designated to coordinate this effort: the Indian Institute of Chemical Biology (CSIR-IICB) for the East zone; the National Chemical Laboratory (CSIR-NCL) for the West zone; the Institute of Genomics and Integrative Biology (CSIR-IGIB) for the North zone and the Centre for Cellular and Molecular Biology (CSIR-CCMB) for the South zone (Figure 1).

### Inclusion/Exclusion criteria

All CSIR permanent staff, including employees, retirees, and their spouses, were eligible for enrollment in this study. The exclusion criteria included pregnancy and being under 18 years of age. Additionally, participants were excluded from a specific scanning modality if they reported a history of medical surgery or complications related to that modality, as detailed in Supplementary Table S2. Participants were instructed to fast for at least 10-12 hours before arriving at the phlebotomy sampling site.

### Tests and scans conducted

Three main categories of assays were planned: blood-based, scanning-based and stool sample-based. Statistically with the objectives defined, we required 3300 subjects as listed in Table 1. The sample size for risk calculation was planned for a regression model with nearly 20 parameters and a c-statistic of 0.85, which was determined with STATA 15 (22-25). Transparent reporting of a multivariable prediction model for individual prognosis or diagnosis (TRIPOD): the TRIPOD Statement and The Strengthening the Reporting of Observational Studies in Epidemiology (STROBE) guidelines for cohort studies will be adhered to, as and when to be reported (26, 27).

**Table 1.** Calculation of Statistical Sample size.

|  | Diabetes |  | Fatty Liver |
| --- | --- | --- | --- |
|  | Q1 | Q2 | Q1 |
| Definition of End Point | What percentage of normoglycaemic population turned diabetic | What percentage of Normoglycaemic population turned pre-diabetic | What percentage of people with fo or f1 fibrosis grading went to f3 or f4 |
| Criteria 1 | HbA1c >= 6.5 from <5.7 | Hba1c btwn 5.7 and 6.5 from <5.7 | Transient elastography reading <6.5 to >=8.6 or 10 |
| Study Design | Longitudinal | Longitudinal | Longitudinal |
| Current Prevalence | 10 | 10.3 | 15 |
| C Statistics | 0.85 | 0.85 | 0.85 |
| Dropout | 30 | 30 | 30 |
| Prediction Parameters | 30 | 20 | 20 |
| Sample Size | 3227 | 1446 | 1074 |
| **Events Needed** | 323 | 188 | 161 |
| **Incidence** | 20 | 13 |  |
| **Years of Follow up** | 3-4 years | 2-3 years |  |

In response to long-term objectives, we could observe that we need ∼3300 samples for events to occur. However, to account for sample availability, considering geographical region, age, sex, etc, and dropout with available resources, we will do phenotyping of 10000 and deep phenotyping of 5000 subjects. It may be noted that the samples of all 10000 individuals will be bio-banked, and if required, based on outcome data, we can do the deep phenotyping on these as well.

A brief description of the test and scans performed and planned is given in Table 2. Biochemical tests and molecular assays, including metabolomics, proteomics, GSA, Cytokine, and Immuno-phenotyping, would be done in a restricted number of samples and not all as described above

**Table 2:** Different assays planned in the Phenome India Project.

|  |  |
| --- | --- |
| **Blood-based** |  |
| 1. Biochemical Analysis | 5. Genomics-Global Screening Array (GSA)*, Whole Genome Sequencing\$ and Whole Exome sequencing\$ |
| 2. Proteomics and Metabolomics | 6. Metal Content Analysis |
| 3. Cytokine* and Immuno-phenotyping*# | 7. Telomere Length and Activity*# |
| 4. RNA-Micro RNA profiling and RNA sequencing# |  |
| **Scanning-based** |  |
| 1. Transient Elastography (TE) for Liver-Fibroscan based | 5. Anthropometry-Height, Dynamometer for grip strength, and anthropometry tape-based measurements of Chest, Waist, and Hip Circumference |
| 2. Spirometry for Lung | 6. ECG |
| 3. Oscillometry for Lung | 7. Skin Health Analysis*\$ |
| 4. Body Composition Analysis (BCA) |  |
| **Stool Sample-based** |  |
| 1. Gut Microbiome sequencing |  |

\*Baseline only, #Subset of samples only,
\$Exploratory

### Cohort enrolment and sample collection

The Institutional Human Ethics Committee (IHEC) approved the study at CSIR-IGIB (reference number: CSIR-IGIB/IHEC/2023-24/16). The study was registered vide CTRI number-CTRI/2024/01/061807. The baseline sample collection was planned for seven months, from December 2023 to June 2024. In each lab, on average, a week was spent sampling nearly 50 participants per day. The two follow-up samplings are planned to be carried out in 2025-26 and 2026-27.

To maximize participation, awareness of the program’s significance and its participatory nature was promoted at each of the constituent laboratories of CSIR through brochures, web-based communications and personal interactions. A signed physical informed consent form was obtained from the participants during sample collection.

### Registration & Survey Questionnaire, Slot Booking

Once awareness of the project was raised within a laboratory, participants were registered through a secure online portal-the CSIR Cohort portal (available at https://www.csircohort.org/) inspired by HIPAA guidelines, the portal implements security measures like two-factor authentication, data encryption, and user timeouts, role-based access, data integrity controls and logging privileges. It also provides supporting documents to inform participants about the study’s objectives and protocols. Using two-factor authentication, participants can access their recorded health parameters and reports anytime through the portal.

Participants completed a baseline questionnaire, which included questions about their family and residence. This information was recorded solely for authentication purposes and stored securely in a separate, encrypted database. Each entry is indexed with a unique Registration ID to ensure participant anonymity.

Each laboratory had a coordinator and a co-coordinator, who were and will be responsible for verifying the authenticity of participants. Once the respective coordinator approves a participant’s registration, they can proceed to fill out the online questionnaire, which is integrated with a secure Research Electronic Data Capture (REDCap) web application hosted in-house for easy self-reporting (28, 29). REDCap is designed to support de-identified data capture for research studies, providing an intuitive interface for validated data entry. Details of the questionnaire are provided in Supplementary Tables S3 and S4.

Participants could only book a slot for sample collection after successfully submitting the questionnaire. They were then able to choose a date and time for their sample collection. Registered participants received notifications about available sample collection dates at their respective labs, allowing them to schedule their visit at their convenience. For microbiome analysis, participants were required to collect the sampling kit within 48 hours of their chosen sampling time. Reminders were sent via email and mobile through the portal. Reports of the anthropometric parameters, scanning-based tests and basic biochemistry were sent to the participants through the portal, which also requires two-factor authentication for access. On the day of the sampling, the participants were provided with a lab ID to maintain confidentiality. The lab ID was generated by the computer against the registration ID provided by the participant.

### Data Acquisition

Each of the activity stations was staffed by trained personnel. All the labs and center stations were equipped with the same equipment except Oscillometry. Details of the various scanning modalities are provided in Supplementary Table S5, and a brief description is provided below. Hard copies of the health card after recording their anthropometric parameters, spirometry, oscillometry, liver elastography results, body composition analysis and ECG were given to the participants. At the end of each day, the station administrator uploads all data in PDF and Excel formats to the portal.

### Anthropometry

Anthropometry was carried out to measure height, weight, chest (CC), waist (WC), abdominal (AC), and hip circumferences (HC). Height (in cm) – As recommended by Integrated Child Development Services (ICDS) guidelines, a standard stadiometer was utilized at all centers. Weight (in kg) - Weight was measured using the Accuniq Body Composition Analyzer (Manufacturer-SELVAS Healthcare, Korea Model BC 380). Body Circumferences (in cm) - CESCORF measuring tape (Manufacturer – Cescorf, Brazil) was utilized to measure CC, WC, AC, and HC. The following definitions were adopted to measure circumferences thrice at each site (30, 31):

CC-“Measure the chest circumference at the most significant part of the chest, which is usually across the level of the nipple line in males and just above the breast tissue in females. Measurements were taken at the end of a normal expiration”.

WC-“A horizontal measure is taken at the midpoint between the lower margin of the last palpable rib and the top of the iliac crest”.

AC-“The tape is held behind the participant with one edge at the horizontal plane through the center of the umbilicus”.

HC-“The participant is standing erect, and the feet close together. A horizontal measure is taken around the widest portion of the hips and buttocks”.

### Dynamometer

It was carried out to record grip strength and the participant was required to sit upright in a chair with feet flat on the floor and legs uncrossed. 200 lbs/90 kg capacity Model 12-0241 Brand-Baseline, USA dynamometer was utilized in the study and was ensured for factory calibration. After positioning the participant comfortably, they would hold the hand dynamometer with their fingers wrapped around one side of the handle and thumb around the other side with the dynamometer set in the second position. The dynamometer should face forward and the forearm should be held parallel to the floor. Participants’ position should be seated with their shoulders adducted, sitting upright with their elbows bent at a 90-degree angle and their wrists set between 0-30 degrees extension. The participant was told to squeeze the hand dynamometer with maximum isometric effort (as hard as possible) for at least 3 seconds and relax. Starting with the right hand, the test was repeated with the opposite left hand and vice versa if the dominant hand was left three times in each hand. It was reset in each repeat. Dynamometer data can be influenced by the duration each participant gets to press it and familiarization with the device. To address these factors, each respondent was given a brief opportunity to acquaint themselves with the dynamometer through a short hands-on practice session. During the test, the dynamometer shouldn’t touch any body part or any other object (32-34).

### Body Composition Analysis

Body Composition Analysis (BCA) was done on the Accuniq Body Composition Analyzer (manufacturer-SELVAS Healthcare, Korea Model BC 380), a three-frequency system and was carried out as per the standard protocol specified by the manufacturer.

### Transient Elastography of Liver

The FS Mini 430 Plus model (Manufacture-Echosens, France) with M or XL probes was used to acquire the scan and data. Fibroscan is a simple and safe technique to assess liver stiffness and fat content, and it takes 5-10 minutes on average per examination. Preliminary preparation required the subject to remain fasting for at least 6 hours before the procedure, which was met through the fasting condition specified. The subject is then asked to lie supine with the right arm in full abduction.

The examiner places the probe along the 5^th^ to 7^th^ intercostal space in the mid or anterior axillary line to obtain a view of the right lobe of the liver. Once the area is identified, measurements are obtained using the appropriate fibro scan probe.

Liver Stiffness Measurement (LSM) and Continuous Attenuation Parameter (CAP) measurements were captured by trained personnel blinded to participants’ clinical data. Although the literature points out the use of an XL probe for body mass index (BMI) greater than 30, we used the recommender system of the device for probe selection. Failure of transient elastography is defined as the inability to obtain valid LSM or CAP value (35-37). Manufacturer guidelines were followed to acquire 10 valid measurements.

### Spirometry

EasyOne Air hand-held spirometer (Manufacturer-NDD, Switzerland), a widely utilized handheld spirometer based on ultrasound sensing, was used in the study to capture spirometry data. American Thoracic Society (ATS)/European Respiratory Society (ERS) guidelines and interpretation were followed to capture spirometry data (38-41). Forced expiratory volume in first second (FEV1) and forced vital capacity (FVC) are generally used to diagnose lung function abnormalities, with an FEV1/FVC ratio of <0.7 indicative of obstructive airway disease (OAD). If the ratio is normal, i.e. ≥0.7, FEV1 and/or FVC of <80% predicted indicate low lung function with restricted lung function measured through FVC and preserved ratio through FEV1 (42).

### Oscillometry

Oscillometry was performed before spirometry using the Tremoflo C-100 Airwave Oscillometry System (Thorasys, Canada) or Resmon Pro V3 System. Oscillometry is used to measure perturbations to sound signals of different frequencies superimposed onto normal tidal breathing, which helps characterize the resistance of small airways and airway distensibility (43). The signal was recorded for at least three artifact-free breaths, which sets the QC before capturing acceptable maneuvers. Resistance and Reactance at 5 Hz (R5 and X5), Resistance at 20 Hz or 19 Hz (R20/R19), Resistance of small airways, 5 to 20 Hz (R5-20 or R5-R19) and frequency response (Fres) were primarily monitored.

### ECG

ECG was performed at a 500 Hz sampling rate using Pisces 1012 model 12 lead ECG, (Manufacturer Allengers, India). The ECG uses a 12-lead setup with a high-pass frequency filter at 0.05 Hz, a low-pass filter at 35 Hz, and a 50 Hz notch filter. The sweep speed was 25 mm/s, and the amplitude was 1.00 cm/mV. A two-minute recording was taken while the participant remained motionless. The data is saved automatically, generating a PDF uploaded to the participant’s dashboard.

### Skin analysis and phenotyping

To develop standards for objective assessment of skin barrier in Indian skin types, skin barrier function was measured by utilizing Courage Khazaka (Manufacturer Courage Khazaka Electronic GmBH, Germany) in terms of four biophysical parameters, employing distinct probes for measurement: Skin sebum content (Sebumeter): The measurement process involves using grease spot photometry with the Sebumeter® SM 815. The matte tape contacts the skin or hair, becoming transparent based on surface sebum. A photocell measures transparency when the tape is inserted into the device aperture, indicating sebum content by light transmission. The Sebumeter measures sebum levels on the forehead and left cheek, with readings taken over 30 seconds at each site. Transepidermal Water Loss (TEWL) (Tewameter): The Tewameter® probe measures the density gradient of the water evaporation from the skin (Δc) proportional to the TEWL. The probe takes three 10-second measurements on the forehead, left cheek, and left volar forearm to assess TEWL. Stratum Corneum Hydration (Corneometer®): The measurement is based on capacitance measurement of a dielectric medium, here the stratum corneum, the uppermost layer of the skin. With increasing hydration, its dielectric properties change. The measurement is based on the fact that water has a higher dielectric constant than most other substances (mainly <7) and is done on the forehead, left cheek, and left volar forearm. Skin Elasticity (Cutometer): The measuring principle of the Cutometer® is based on the suction method, where negative pressure deforms the skin mechanically. The device creates pressure, drawing the skin into the probe’s aperture. After a defined time, it is released again.

### Blood Collection, Testing and Storage

The protocol for blood sample collection, preparation and management was conducted in collaboration with a pathology laboratory that ensured the collection of high-quality samples for subsequent research purposes. The routine blood tests described in Supplementary Table S6 were performed by the diagnostic laboratory, which was accredited by the National Accreditation Board for Testing and Calibration Laboratories (NABL) and the College of American Pathologists (CAP). Other biochemical analyses are being performed at CSIR-IGIB (Supplementary Table S7). The results are uploaded to the central database, and the participants are provided passcode-enabled access to their results. The data is stored in a data repository at CSIR-IGIB with the provision of a backup at CSIR-4PI. The sample flow and processing work plan is depicted in Figure 2.

**Figure 2:**
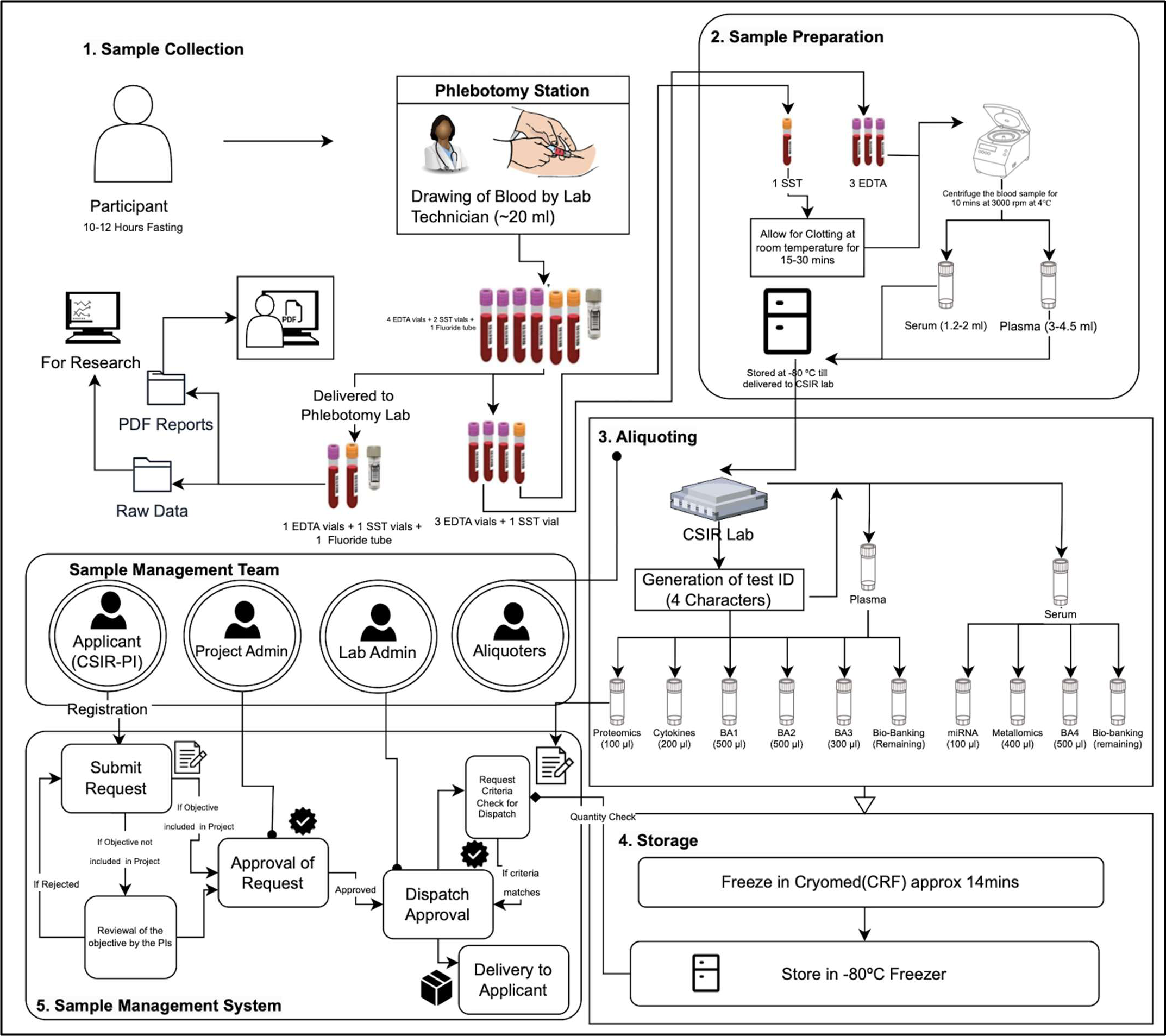
Sample flow and processing in Phenome India Project.

### Blood Sample Collection

Participants came to the collection centers with prior fasting for 10-12 hours and 20 mL of blood was drawn at the phlebotomy station into specific blood tubes such as Ethylenediamine tetraacetic acid (EDTA) (4 tubes), serum-separating tube (SST; 2 tubes), and Fluoride (1 tube). (One EDTA, One-Serum and One Fluoride tube were transported, maintaining a proper cold chain to the pathology laboratory for basic biochemistry analysis. Plasma and serum samples separated in the sample collection site were transported using a cold chain to CSIR-IGIB, where they were aliquoted, provided a test ID and stored at -80°C for future analysis. The sample flow and processing work plan is shown in Figure 2.

### Bio-Banking

Each aliquot was assigned a unique 4-character Test ID and frozen using a Cryomed Controlled-Rate Freezer (CRF) for approximately 14 minutes before being stored in an ultra-low deep freezer. The samples are stored in 0.5 ml 2D-tubes marked with unique quick response (QR) codes and tube numbers for precise traceability and data confidentiality. A robust sample management system is maintained at the laboratory that can track samples and facilitate their future usage. A meticulous biobank system will ensure the expansion of global biobank networks and comprehensive data-sharing frameworks to enhance the scope and impact of research. It will further act as a foundation for reliable research by accurately correlating samples with participants and health outcomes.

### Assays to be performed and planned

The different experimental assays to be performed and planned are summarized in Table 3.

**Table 3:**
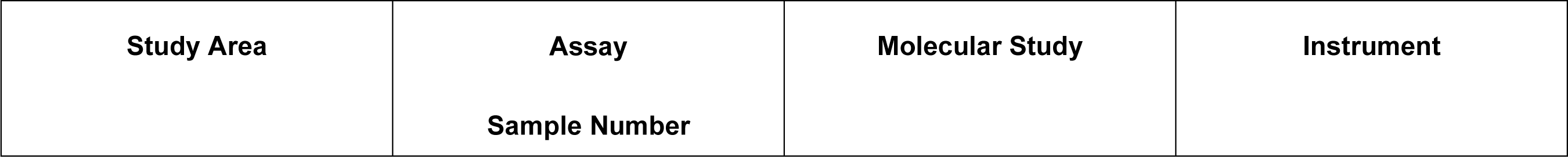
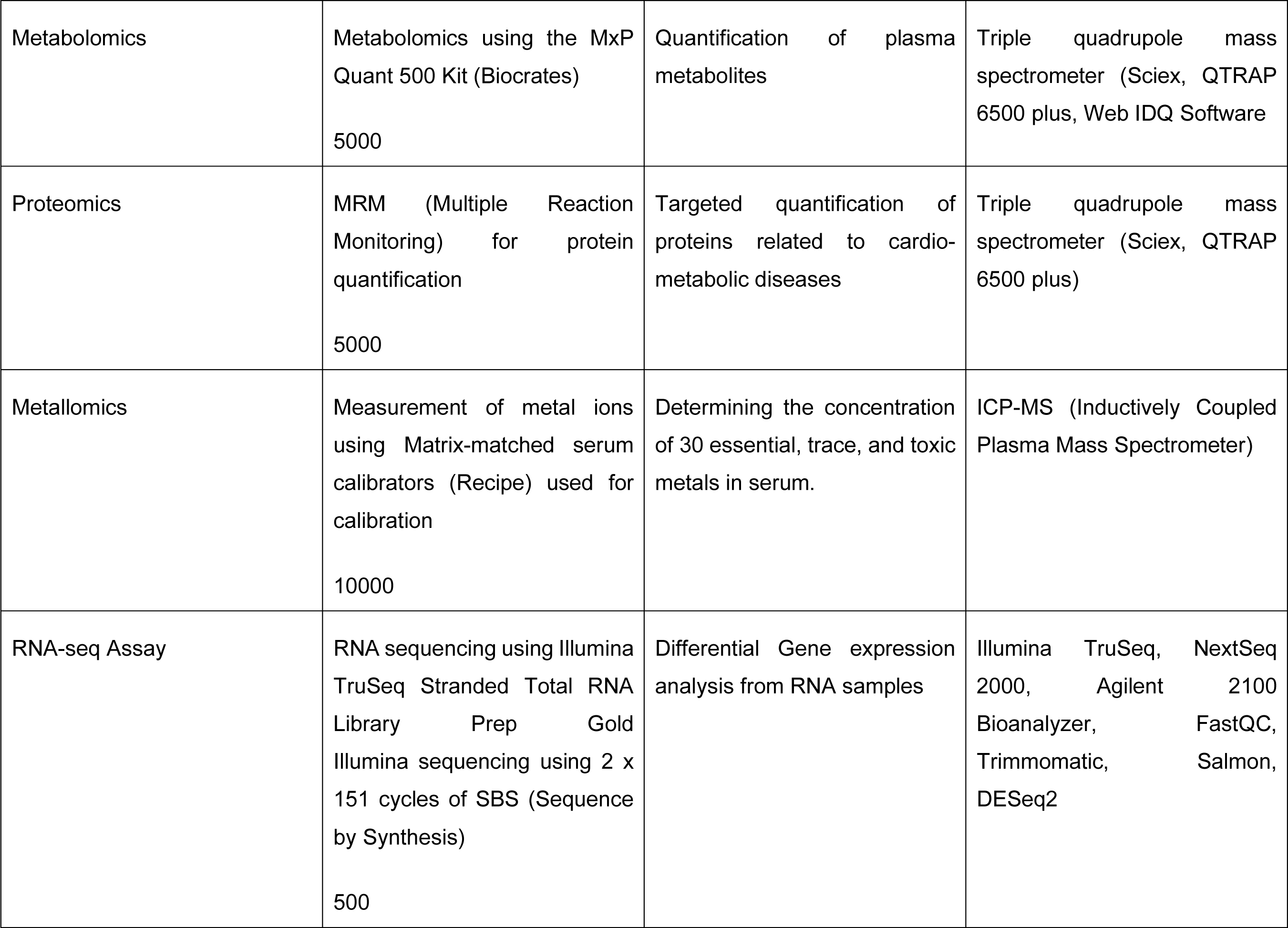
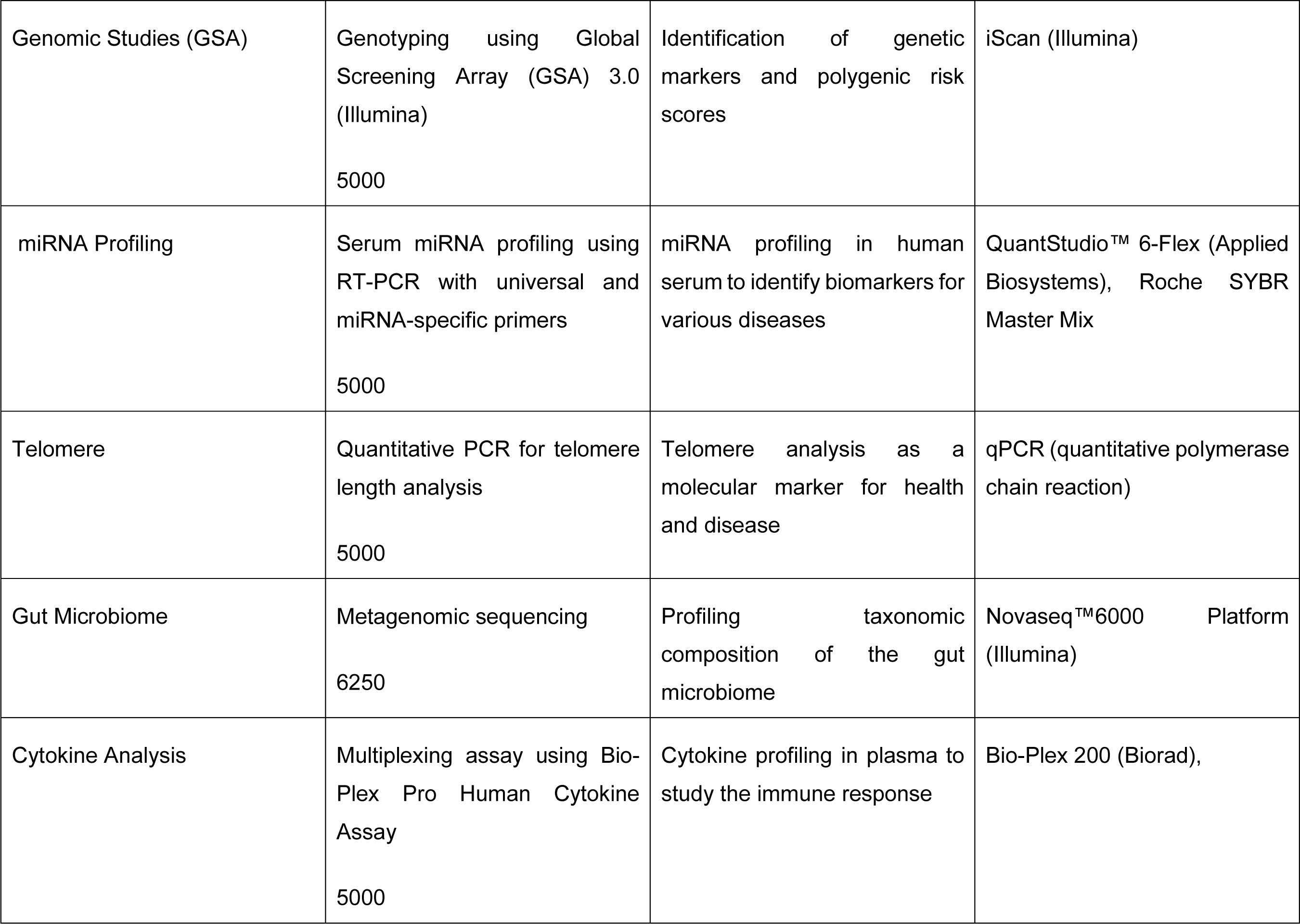
Summary of the different assays proposed in the Phenome India Project.

| Study Area | Assay<br>Sample Number | Molecular Study | Instrument |
| --- | --- | --- | --- |
| Metabolomics | Metabolomics using the MxP Quant 500 Kit (Biocrates)<br>5000 | Quantification of plasma metabolites | Triple quadrupole mass spectrometer (Sciex, QTRAP 6500 plus, Web IDQ Software) |
| Proteomics | MRM (Multiple Reaction Monitoring) for protein quantification<br>5000 | Targeted quantification of proteins related to cardio-metabolic diseases | Triple quadrupole mass spectrometer (Sciex, QTRAP 6500 plus) |
| Metallomics | Measurement of metal ions using Matrix-matched serum calibrators (Recipe) used for calibration<br>10000 | Determining the concentration of 30 essential, trace, and toxic metals in serum. | ICP-MS (Inductively Coupled Plasma Mass Spectrometer) |
| RNA-seq Assay | RNA sequencing using Illumina TruSeq Stranded Total RNA Library Prep Gold<br>Illumina sequencing using 2 x 151 cycles of SBS (Sequence by Synthesis)<br>500 | Differential Gene expression analysis from RNA samples | Illumina TruSeq, NextSeq 2000, Agilent 2100 Bioanalyzer, FastQC, Trimmomatic, Salmon, DESeq2 |
| Genomic Studies (GSA) | Genotyping using Global Screening Array (GSA) 3.0 (Illumina)<br>5000 | Identification of genetic markers and polygenic risk scores | iScan (Illumina) |
| miRNA Profiling | Serum miRNA profiling using RT-PCR with universal and miRNA-specific primers<br>5000 | miRNA profiling in human serum to identify biomarkers for various diseases | QuantStudio™ 6-Flex (Applied Biosystems), Roche SYBR Master Mix |
| Telomere | Quantitative PCR for telomere length analysis<br>5000 | Telomere analysis as a molecular marker for health and disease | qPCR (quantitative polymerase chain reaction) |
| Gut Microbiome | Metagenomic sequencing<br>6250 | Profiling taxonomic composition of the gut microbiome | Novaseq™6000 Platform (Illumina) |
| Cytokine Analysis | Multiplexing assay using Bio-Plex Pro Human Cytokine Assay<br>5000 | Cytokine profiling in plasma to study the immune response | Bio-Plex 200 (Biorad), |

## Data acquisition and storage

### Data Management

The backend application programming interface (API) and frontend application were integrated and deployed primarily on a virtual machine at CSIR-IGIB. Data was stored in a relational database management system (RDBMS)-based architecture on the back end. The front end was developed using server-side scripting language-based architecture and framework with modular supports from JavaScript, CSS (Cascading Style Sheets), and web server-related programming widgets. Robust data security and access management protocols were deployed, and development using non-standard protocols was avoided to prevent potential SQL (Structured Query Language) Injection attacks. Data backup, recovery, and mirrored deployment (disaster management protocols) are proposed (Figure 3).

**Figure 3:**
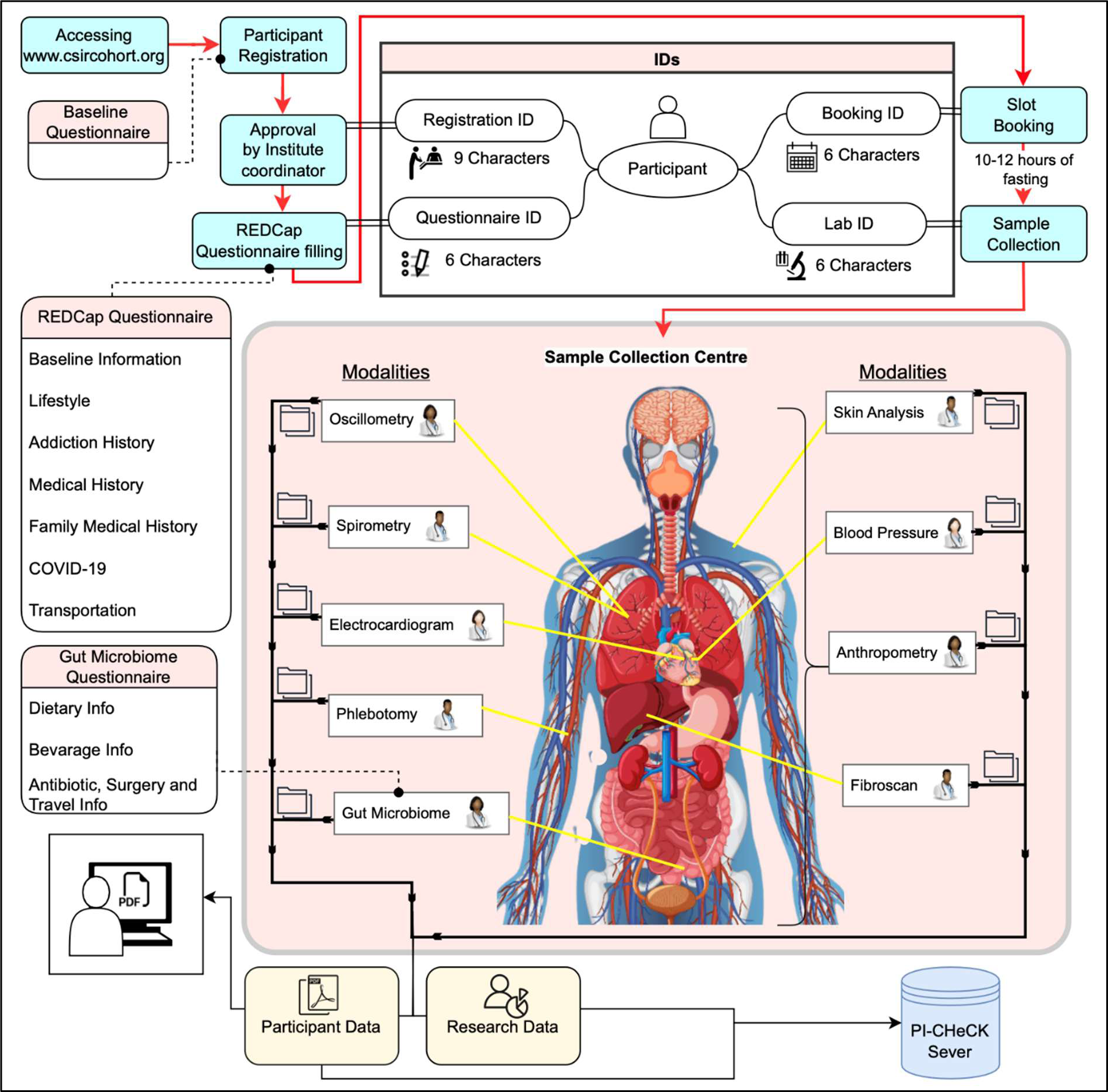
Overall flow of data in the Phenome India Project.

### Data acquisition

The data acquisition process for the PI-CHeCK study involves collecting data from 10,000 participants of diverse backgrounds initially over a five-year period. The study includes surveys, laboratory tests, clinical tests, and scanning modalities. Data is uploaded via a secure protocol involving restricted access to the station admins of the respective modality. To protect data security and privacy, participant information is de-identified, and Base64 encoded, with one-time password-enabled restricted access granted only to authorized personnel. The data is stored in the in-house server, established at CSIR-IGIB, where the PI-CHeCK portal is hosted. The data backup is planned to be managed by a scheduled task that automatically stores backups on the same server and a remote server (through file transfer protocol) at regular intervals. Researchers may need access to participants’ data stored in a centralized database for their studies. However, access to this data is controlled to ensure participant privacy and compliance with regulations inspired by health insurance portability and accountability act (HIPAA) guidelines.

### Data security and privacy

Apart from basic security measures of password hashing, one-time password-enabled access and security headers, the portal is equipped with security measures to counter cross-site request forgery and handle injection attacks by implementing security cross-site scripting. Arrangements are made to encrypt data in transit and rest in a secure server. Further, the management of sessions strategically prevents interception and unauthorized access.

Utmost care has been taken to maintain data privacy, i.e., data is secured at rest, during transit, and editing. Collected multi-omics/clinical/lifestyle data from different CSIR institutes is stored in a relational database management system. The database can be queried using a secured graphical user interface (GUI) application. Individuals (who provided samples) were treated as ‘normal users’ of the application and allowed only to view the submitted data. Power users with ‘Lab’ privileges can generate sample IDs (multiple samples for each ‘normal user’), add/link protocols, and upload data files (raw and processed) against anonymized sample IDs only.

A centralized data platform capable of encompassing about 500 terabytes (TB) and appropriate data security, including data protection and database development, was set up at CSIR-IGIB. CSIR-4PI has been recognized as a data recovery site. To enable collaboration amongst the participating CSIR labs, curated de-identified data will be made available for in-depth analysis.

### Data Sharing

Data accessibility is enabled for authorized users or systems to access specific datasets. It involves establishing protocols and permissions to ensure that only authorized users can access the necessary data. Access to participants’ data is strictly controlled to ensure privacy and regulatory compliance. Authorized researchers with approved projects must submit a Data Access Request Form, reviewed by a Data Access Committee to avoid overlapping objectives. Access is granted for a defined period, with data sharing restricted to individuals listed in the request. Role-based access control (RBAC), encryption, and audit logs are implemented to prevent unauthorized access and track data usage. Secure data sharing using secure file transfer protocol (SFTP) is essential for maintaining sensitive research data’s confidentiality, integrity, and availability, ensuring compliance with ethical and regulatory standards. SFTP provides a secure alternative to traditional methods such as email and FTP by encrypting data during transmission and implementing robust authentication mechanisms. Our protocol includes configuring the SFTP server, establishing user groups with appropriate permissions, and setting up secure directories, with additional measures like virtual private network (VPN) configuration and tools such as graphical user interface or command-line interfaces (CLI) to enhance security and efficiency.

### Data backup and mirroring

Data backup and mirroring efforts will be implemented at the CSIR-4PI facility situated in the south zone, which is geographically distinct from the primary data storage site in the north zone.

### Data Analysis

In addition to data visualization, we plan to analyze inter-parameter linear/nonlinear correlation keeping given age/gender covariates, use open source tools to identify patterns within the data, develop predictive models using machine learning (ML) algorithms to predict certain phenotypic conditions, create an integrative network of various genomics, proteomics, and metabolomics parameters to understand the systemic regulation (Figure 4).

**Figure 4:**
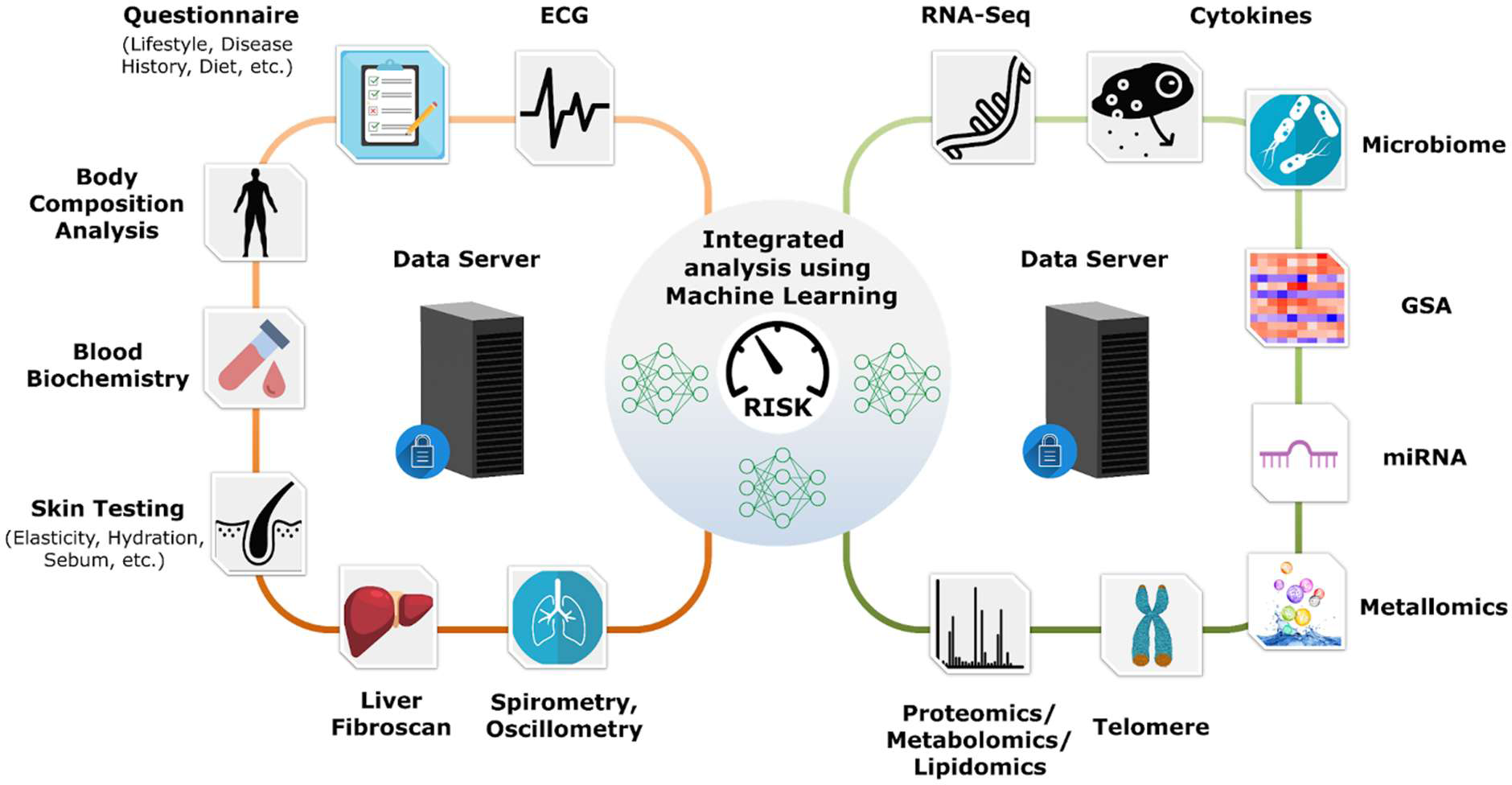
Schematics depicting multi-modal data integration for risk score development.

Sample and indexed participant similarity can be achieved through unsupervised approaches including, but not limited to, clustering and Euclidean/Mahalanobis distance-based methods, which can be used to quantify these similarities. These similarity scores can then be fed and used to train personalized models for risk score assessment.

Using collaborative filtering techniques, individuals having similar phenotypes can be clubbed together. Employing cohort participants’ similarities will help identify a subset proximal to an index patient, which will then be used to train a personalized model in a disease-specific manner. The approach for the personalized model will depend on the type of disease and ground truth label criteria. Recent studies have used various algorithms ranging from recurrent neural networks and ensemble techniques as personalized models for disease state prediction (8, 9). Multi-view datasets will allow us to explore the quantitative association of collected risk factors with the comorbidity- and disease-specific outcomes. The developed framework will be based on explainable AI, rendering the weight of each contributing factor to the predicted outcome.

## Discussion

The incidences of metabolic and other non-communicable complex diseases are increasing at an alarming rate in India which is now considered to be the hub of cardio-metabolic diseases and diabetes in the world (19, 44). From the public health perspective, it is of prime importance to understand the risk factors driving this rise in disease prevalence and develop effective risk stratification, prevention and management strategies.

Large-scale longitudinal cohort studies are essential for investigating disease causation and linking risk factors to health outcomes. For example, the Framingham Heart Study, an ongoing study initiated in 1948, has provided valuable insights into risk factors for heart disease, such as lipid levels, diet, exercise, hypertension, cigarette smoking, etc. This research led to the development of the Framingham Risk Score (FRS), a 10-year cardiovascular risk prediction tool (45-49). However, the FRS may be less applicable to Indian patients due to diverse lifestyles and risk profiles, highlighting the need for a tailored risk matrix based on long-term data specific to the Indian population (50, 51). Over the last few decades, several studies have aimed to identify diagnostic and prognostic markers in India. Most of these were case-control studies, focused on a limited number of biochemical markers and lacked the capacity to analyze critical disease trajectories (52-56). Therefore, a comprehensive longitudinal study across India is necessary to explore the complex interactions between environmental, genetic, and molecular factors influencing disease risk.

The Council of Scientific and Industrial Research (CSIR), with its extensive network of over 37 laboratories and centers nationwide, is ideally positioned to address these gaps. This capability was demonstrated during the COVID-19 pandemic, where a longitudinal cohort provided vital insights into infection dynamics, antibody response dynamics, herd immunity trends across India, and the likelihood of significant outbreaks in a population (16, 57-59). The study helped obtain sentinel data about the spread pattern and different characteristics of SARS-CoV-2 infection across India.

Recent advances in molecular techniques in genomics, transcriptomics, proteomics, metabolomics, and metagenomics have significantly enhanced our understanding of several facets of human biology. Through the Phenome India project, CSIR aims to leverage these advancements alongside the prospective collection of biological samples from a diverse population, enabling long-term follow-up to establish causal relationships and develop diagnostic and prognostic biomarkers for chronic non-communicable diseases. The extensive phenotyping of participants will make this cohort unique and invaluable. To the best of our knowledge, tests like fibroscan, oscillometry, and skin barrier assessments are being conducted in such a large and diverse population for the first time in India. The prospective collection and analysis of fecal microbiomes adds a new dimension to understanding cardio-metabolic disease risk. Additionally, evaluating metal content in participant’s serum samples together with metabolomics, proteomics and genomics will help illuminate the complex interactions between genetic, dietary, and environmental factors impacting metabolic diseases. This deep phenotyping is also expected to unravel several novel genotypic and phenotypic associations in the Indian population.

Employee-based cohorts and restricting a longitudinal cohort study to urban and semi-urban areas introduce both limitations and advantages that can impact the study’s generalizability and data quality and feasibility. Employees, especially those in formal jobs, may not represent the general population due to differences in socioeconomic status, education, and access to healthcare. Urban and semi-urban populations differ significantly from rural populations in terms of lifestyle, healthcare access, and environmental exposures. Urban and semi-urban populations may have better access to healthcare and technology than the rural populations. The significant socio-demographic differences between urban and rural populations and gene-environment correlations in rural and urban populations may differ in their genetic risks for certain health outcomes and it could bias the study’s findings. This could skew the findings and limit the findings of health trends across different geographic regions.

However, there are advantages to an employee-based cohort focused on urban and semi-urban areas, particularly in terms of recruitment and follow-up feasibility. Educated participants are more likely to understand the study’s importance and comply with data collection and follow-up protocols. Employees are more likely to come from middle or upper socioeconomic backgrounds, which may be seen as potentially skewing the health profile of the cohort and limiting insights into diseases associated with poverty; however, it makes it a uniform cohort and homogenous in terms of socioeconomic strata, education and access to healthcare. The current cohort while it may miss conditions prevalent in rural areas, such as certain infectious diseases, malnutrition, or specific chronic conditions, it would stand as one of a kind for a homogenous population being screened at this large scale and specifically in the context of non-communicable diseases

In conclusion, this multicenter, multi-modal prospective cohort study represents a significant step towards precision medicine, in the Indian context. This level of comprehensive phenotyping has not been attempted before and when combined with data analytics and AI/ML approaches, it aims to produce reliable risk prediction tools for cardio-metabolic disorders.

## Ethics

The study was approved by IHEC at CSIR-IGIB (reference no.: CSIR-IGIB/IHEC/2023-24/16). The study was registered vide Clinical Trials Registry-India (CTRI) number-CTRI/2024/01/061807.

## Data Availability

Anonymized Data for public use may be made available after 3 yrs from completion of baseline phase of study or as per advisory from Monitoring Committee of the project if any revisions thereof.

## Acknowledgments

We acknowledge the support from CSIR, participants, and volunteers. Further, following names are acknowledged:

Arpana Parihar, Shalu Yadav, Mohd Abubakar Sadique, Akshay Singh Tomar, Varsha Agrawal, Sakshi Rajput, Aabha Kushwah, Shilpee Chauhan, Varsha Parmar, Pushpesh Ranjan, Neeraj Kumar, Abhijeet Bamoria, Abhisek Giri, Navnidhi Tripathi, Jyoti Lodhi, Parul Shrivastava, Prachi Shrivastava, Komal Sharma (**AMPRI**), Vikas Pathak (**CBRI**), Surya Narayan Mishra, Sathya Swetha, Gururaj Kalshetti, Rakesh Pal Bhagat, Chandreshwar Raju Kataru, Hari Charan Goud Nerella, Amit Chakraborty, Prangya Paramita Sahoo, Alagu Sankareswaran, Sofia Banu, Priyadarshini Sanyal, Sourav Ganguly, Manash Kumar Behera, Pallavi Rao T, Akila Ramesh, Niharika Tiwary, Devika Mahimakar, Abhishek Saha, Shreya, Pramoda, Ruqaiya Tasneem (**CCMB**), Khushbu Singh Raghav, Shivam Tiwari (**CEERI**), N Vinodkumar, Rani C Avilash Sumithra, Pramila Epparti, M Archana (**CFTRI**), Swachchha Majumdar, Sourja Ghosh (**CGCRI**), Debabrata Chanda, Dnyaneshwar Umrao Bawankule, Pankhuri Singh, Pankaj Kumar Shukla, Parmanand Kumar, Sumati Sen, Chandra Kant, Sarita Upadhyay, Sumit Kushwaha, Sukriti Srivastava, Daneshvar Prasad, Kavita Singh, Om Prakash Choudhary Kar, Debasmita Sahoo, Anant Kumar Chaudhary (**CIMAP**), Srinivetha Pathmanapan, Syed Nasar Rahaman, Vikash Negi, Jegan K, Sankar Palanivel, Lishadevi M, Pranathy K, Vandhana Anumaiya, Kirubakaran BJ, Thiyaneshwar (**CLRI**), Sujata Pachhal, Sukanta Maji, Sonali Garai (**CMERI**),Mukti Advani, Rina Singh (**CRRI**), Sandeep Kumar, Rajni, Preetismita Borah, Manish Kumar, Goraj Singh, Suraj Prakash, Ajay Kumar, Inderjit, Shubham Patial, Shavita, Amandeep, Jyoti Jangra, Aman Grewal, Shivalika, Rajrani, Swapna, Dinesh Kumar, Dhiru, Munish, Akriti, Deepak Kumar, Gourav, Komal, Prashant Kumar, Amrit Marthu, Rishav Rai, Amir Khan, Satinder Kaur, Netra, Pankaj, Anuj Sharma, Parneet Kaur, Krishna, Pallavi Suman, Abhishek, Yash (**CSIO**), Haresh Khasiya, Vishal Dumraliya, Chinmay Chauhan, Priyanka Pandya, Disha Rohit, Neha Sharma, Aadya Yadav, Riya Soni, Dhaval Nathani, Mujeer Habsi, Chand Kothadia, Parth Nathani, Keyur Patel, Jigar Aal, Muskan Kalla, Jayesh Rathod, Saif Syed, Abhit Shah, Chirag Khambhaliya, Mrunalini Gohil, Aditya Vaja, Asmita Dhimmar, Rupal Dubey, Jalak Maniar, Kashish Sharma, Dhruvil Chavda, Charu Waghmare, Anushka Tiwari, Shravi Jain, Arun Rathod, Ankita Shandul, Bharti Rana, Deepesh Khandwal, Tulika Mairal, Rajveer Parmar, Mohit Dangariya (**CSMCRI**), Ajeet Singh (**HRDC**), Salwa Naushin, Subhash Gurjar, Rajesh Kumar (**IGIB**), Anuj, Aditya Ranout, Trilok Saini, Sahdev Choudhary, Amit Kumar, Shweta Sharma, Neha Bhardwaj, Kajal Kalia, Suresh Kumar, Abhishek Goel, Ravi Kumar, Surbhi Mali, Komal Goel, Swati Katoch, Shagun Dogra, Vinesh Sharma, Rajneesh Kumar, Poonam Dhiman, Pardeep Poonia, Sahiba Chahal, Rahul Kumar, Athrinandan s Hegde, Rupinder Kaur, Vivek Kumar, Prakriti Sharma, Avisha Sharma, Ankita, Mohak Mali, Priyanka Koundal, Ashruti (**IHBT**), Rajendra Prasad Punna, Kiran Kumar A, Taslim B Shaikh, Nidhi Sharma, Anjali Veeram, Komal walvekar, Mani Sharma, Suriya panneer, Rajwinder Kaur, Ramasatyasri Kotipalli, Hari Priya Sripadi, Vaishnavi Kambhampati, Pinkal Sondarva, Divya Vani kundurthi, Maneesha Yeddula, Sindoora Dhoolipala (**IICT**), Abhilek K Nautiyal, Harleen Kaur, Ayan Banerjee, Rahul Gautam, Mirnalini Juyal, Mridul Budakoti, Preetanshika Tracy, Pratima Ashoke Patel, Ritu Mourya, Harish Panwar (**IIP**), Sneha Mohanty, Mohd Tauseef Khantwal, Vaibhavi Lahane, Shreya Tripathi, Sunita Devi, Gulafsha Siddiqui, Km Priya, Priyanka Goswami, Sandip Chatterjee, Sakshi Singh, Sachin Mishra, Abhineet Singh, Vishal Narayan Soni, Devendri Khantwal, Murk Kumari, Sanjivani Naik, Apoorva Dewangan, Ananta Joshi, Akansha Chaurashiya, Akansha Chauhan, Sheetal Agarwal, Joel Saji, Nikita Shraogi (**IITR**), Ankita Das, Mayuri Kumari, Amit Kumar Swain, Deepak Kumar Ahirwar, Nilotpal Kapri, Ghrutanjali Sahu, Santosh Kumar Behera, Suchismita Senapati, Subhashree Nayak, Yogesh Chaudhary, Anagonou Bertrand, Bibekananda Nayak, Suraj Kumar Sahu, Saswati Suryasnata, Champakeswara Mahanta, Annapurna Behera, Satyabrata Sahoo, Akshyurna Pattnaik, Rakhi Rani Biswas, Chandrama Dipa, Adyasha Bijay Mishra, Dinesh Kumar Panda, Sudipta Nayak, Avishek Kar, Kajal Sundaray, Swagatika Dash, Alok Pattanaik, Sonali Swapanjali Bhoi, Swikruti Mishra, Shalini Das, Parinita Mishra, Debidatta Barik, Tejaswini Das, Subhashree Pattnaik (**IMMT**), Kailash T Bhamare, Chander Shekhar Sharma, Amit Kumar, Vineet Kumar, Deepak Bhatt, Surjeet Singh, Harminder Singh, Paramjit Lal, Sandeep Kumar, Nitin Sharma, Jaideep Mehta, Renu, Rohtas Ranga, RK Dhiman, Anjali Koundal, Bhumika Vaidya, Amit Kumar, Shweta Pandey, Anunay Sinha, Joyshree Das, Shubham, Bulbul Roy, Pompi Bhadra, Meena Sharma, Sana Khatun, Payal Thakur, Bhagyashree Rabha, Meenal Rastogi, Mayur Sudhakar Zarkar, Somnath Chindhe, Shreya Singh, Neelam (**IMTECH**), Kiran Raj S, Ravi Kumar Sharma Kumar (**IPU**), Nataraj D, Dr. Swetha C Desai, Shivanna TB, Murthy BYK, Raghu G, Sunil A, Shashidhara KN, Raghavendra Swamy M, Aravind Kumar Joseph, Purushotham HM, Shiva Kumar SR, Muniprakash M, Shekhar Gogeri, Siva Kumar R, Jaikesh S, Ranganatha S, SatishKumar Shri, Shashikala U, Praveen Kumar JD, Jitendra Ping (**NAL**), Koteswara K Rao, Parveen Goyal, NMR Ashwin, Rakesh S Joshi, Rupali Waichal, Shashikala Ranjane, Abujunaid Khan, Priyanka M Bankar, Rachel Samson, Apurva R Barge, Ashish S Jagtap, Amreen Sheikh, Shahaji Palaskar, Jugal Kanerkar, Arvind Chourasiya, Bhagyashree Likhitkar, Pradeep S, Deepak Jadhav, Pranay Awathare, Swaraj Jathar, Pooja Deshmukh, Vaishnavi Salunkhe, Bharat Pande, Aruna Choughule, Yugendra Patil, Rohit S Dashpute, Minal R Bhalerao, Vinod Kamble, Yashodhara Shinde, Vineetkumar S Nair, Prathmesh Ghongade, Prashant Kalaskar, Ganesh Jadhav, Sakshi Pingle, Preshitta Bhat, Shubham Choure, Tanaji B Devkate, Vikram Nichit, Shrutika M Shewale, Shyam K Gawari, Sonali Gaikwad, Abhishek Tripathi, Sayli Jamdade, Ashtamy MG, Shubham Kumar, Shivani V Palkar, Rutuja Ugle, Kaumudi Joshi, Khushboo Bavishi, Priya Bhavsar, Nandini Rangari, Ayushi Rushesari, Maria Kuwazerwala, Taha Farhan Siddiqui, Anand Kumar Shukla, Prakashini Saroj Nilgirwar, Sakshi R Mangate, Sindhu Bali, Prabha Oli (**NCL**), Pankaj Pardhi (**NEERI**), Jayashree Chiring Phukon, Shridhar Hiremath, Prachurjya Dutta, Parishmita Borgohain, Moirangthem Goutam Singh, Devpratim Koch, Dipanneeta Das Gupta, Pankaj Barman, Sahana SK, Monojit Kumar Roy, Aditya Sarkar, Bhaben Sharma, Shyamalima Mech, Masum Saikia, Trishna Rani Borah, Gayatri Gogoi, Anupriya Borah, Udeshna Changmai, Himadri Das, Anshuman Goswami, Rocktotpal Mahanta, Rina Yumnam, Sukanya Borthakur, Manabendra Borah, Sarangthem Dinamani Singh, Pronami Gogoi, Priyanka Saikia, Umakanta Tanti, Mohan Kurmi, Ravi Kumar Sahu, Nayan Jyoti Borah, Babli Borah, Sindhu Sharma, Trishna Dutta, Sarifa Mafuz Ahmed, Bidyut Prakash Deka, Darshana Tamuli, Meghna Kakoty, Bhaswat V. Borah, Yashodhara Goswami, Priyanka Boro (**NEIST**), Inshamol KP, Athulya, Akhila, Amrutha M, Anaga, Anaswara P A, Anirudh, Anjitha, Anupama, Anusha, Anusree P, Aparna, Aparna Dinil, Arathy, Archa R S, Archana, Arnold, Athira V C, Biji Raphy, Dileep R Nair, Evan, Gayathri, Gopika R, Greeshma, Greeshma Jayan, Karthika N, Karthika Nath, Kavya Mohan, Krishan Unni, Lakshmi M Nair, Lakshmi Shaji, Nandana A B, Neeraja, Neeraja M, Neetha P Sobandas, Parvathy, Reena R, Saranyadevi, Shahansha, Shamna Fathima, Shehbas C, Shilpa, Sruthi, Swapna B, Varsha, Vishnu, Valan Rebinro, Adarsh VP, Jedy Jose (**NIIST**), Govind Ranade, AS Unnikrishnan, Sohan Pal Meena, Karishma Pradeep Chari, Vitasta Jad, Vikash Kumar, Nishamol M, Helen Agnes, Natasha Maria Barnes, Bhumika Rohidas Shirodkar, Shania Waluscha Moeres, Yuvrani Halarnkar, Ramila Ram Gaonkar, Mrinalini Chandra Mohan, Pratibha Bachhley (**NIO**), Himani Meena, Abhinav Banait, Shekar Sharma (**NIScPR**), Ashok Kr Shaw, T Kr Minz, Rahul Tirkey, Mousumi Sarkar, Chandan Kumar Chowdhury, Raushan Kumar, Amit Kumar, Sudip Kumar, Surajit Kundu, Mahesh Pramanik, Deepak Dhivar, Aman Beldar, Tapan Kumar Shaw, Premlal, Akul Gope, Nisha Gupta, Alka Kumari, Vinod, Shikha Kumari, Abhishek Kumar Machhua, Uttam Mukhi, Rahul Mukhi, Manju, Jaydev Sahu, Ravi Ranjan Kumar (**NML**), Rajesh, Sumana Gajjala, Sudesh Yadav, Vinod Kumar Tanwar, Vishesh Garg, Manoj Kumar Pandey, Vikash Sharma (**NPL**)

## Author Contributions

## Phenome India Consortium Authors

## Project conceptualization

Shantanu Sengupta, Debasis Dash, Viren Sardana, Kumardeep Chaudhary (**IGIB**), Giriraj Ratan Chandak, Swasti Raychaudhuri, Karthik Bharadwaj Tallapaka (**CCMB**), Mahesh J Kulkarni (**NCL**), Partha Chakraborty, Dipyaman Ganguly (**IICB**), Umakanta Subudhi (**IMMT**).

## Project planning/Collection management

Shantanu Sengupta, Debasis Dash, Viren Sardana, Kumardeep Chaudhary, Vamsi K. Yenamandra, Rakesh Sharma, Aastha Mishra, Ajay Pratap Singh, Swarnendu Bag, Beena Pillai, Vivek Rao, Sheetal Gandotra, Shantanu Chowdhury, Rajesh Pandey, Bhavana Prasher, Pankaj Pandey, Ankita Sahu, Sudhir Rohilla, Komal Jindal, Vivek Junghare, Tarani Mathur, Meghana Arvind, Satyartha Prakash, Yogesh Kumar, Vikas M Hiremath, Vignesh S Kumar, Deeksha Yadav, Deepak (**IGIB**), Swasti Raychaudhuri, Giriraj Ratan Chandak, Karthik Bharadwaj Tallapaka, Rakhesh K V (**CCMB**), Ashok P Giri, Narendra Y Kadoo, Mahesh J Kulkarni, Dhanasekaran Shanmugam, Mahesh S Dharne, Syed G Dastager, Chiranjit Chowdhury (**NCL**), Partha Chakraborty, Dipyaman Ganguly, Arun Bandyopadhyay, Shilpak Chatterjee (**IICB**), Umakanta Subudhi, Bhavani S Jena, Trupti Das, Boopathy Ramasamy, Pavan Kumar (**IMMT**), Ramakrishna Sistla, Prabhakar Sripadi, Jagadeshwar Reddy Thota, Ramesh Ummanni, Srinivasa Rao M, Sai Balaji Andugulapati (**IICT**), Amit Lahiri, Mrigank Srivastava, Vivek Bhosale (**CDRI**), Iranna Gogeri (**4PI**), Raju Khan, Narendra Singh (**AMPRI**), Rajesh Kumar Verma, Neeraj Jain (**CBRI**), Ganesh Venkatachalam, Murugan Veerapandian (**CECRI**), Deepak Bansal, Amit Kumar, Dinesh Gupta(**CEERI**), Prakash M Halami, S P Muthukumar (**CFTRI**), Anirban Pal, Anil Kumar Maurya (**CIMAP**), Jai Krishna Pandey, Bhanu Pandey, A K Raman (**CIMFR**), Suresh Kumar Anandasadagopan (**CLRI**), Swati Saha, Vishal Anand (**CMERI**), Suman Singh, Anamika Kothari (**CSIO**), Avinash Mishra, Mangal S Rathore (**CSMCRI**), Preeti Srivastava, Pooja Aggarwal (**HQ**), Shreedhar Kanagarjan (**HRDG**), Yogendra Padwad, Vikram Patial (**IHBT**), Sumit G Gandhi (**IIIM, Jammu**), Fayaz Malik (**IIIM, Sinagar**), Debashish Ghosh, Jyoti Porwal (**IIP**), Vikas Srivastava, Prabhanshu Tripathi (**IITR**), Anshu Bhardwaj, Srinivasan Krishnamurthi, Rashmi Kumar, Deepak Sharma, Amit Tuli (**IMTECH**), Indrani Ghosh (**IPU**), Prakash L, Satisha Shri (**NAL**), Chandana Venkateswara Rao, Sanjeev Kumar Ojha, Brahma Nanda Singh, Vijayanandraj Selvaraj (**NBRI**), Shilpa Paranjape, Prashanti Niwant (**NEERI**), Jatin Kalita, Prasenjit Manna, Romi Wahengbam, Tridip Phukan, Pankaj Bharali (**NEIST**), E V S S K Babu, Biswajit Mandal, T Vijaya Kumar (**NGRI**), Rajeev K Sukumaran, Rameshkumar N (**NIIST**), Samir Ravikant Damare, Sameer Damare,Kalpana Sandesh Chodankar (**NIO**), Arvind Meena, Arun Uniyal (**NIScPR**), Ansu J Kailath, Krishna Kumar, Roshan Kumar, Nikhil Kumar, Kuldeep Singh Gour (**NML**), Ved Varun Agrawal, Arun Kant Singh (**NPL**), Maheswaran Srinivasan, Vasudevan Pandurangan (**SERC**), Lalita Goyal (**TKDL**), Rashmi Arya, Manisha Sakpal (**URDIP**).

## Portal development and management

Kumardeep Chaudhary, Debasis Dash, Viren Sardana, Ajay Pratap Singh, Shantanu Sengupta, Satyartha Prakash, Vignesh S Kumar, Anshul Verma, Safeer Khan, Anshul Bhardwaj, Sudhir Rohilla, Prateek Singh (**IGIB**), Gopal Krishna Patra (**4PI**), Nikhilesh Yadav (**NCL**), Abbani Rakesh (**NAL**)

## Sample Collection / Scanning

Pankaj Pandey, Sudhir Rohilla, Vivek Junghare, Komal Jindal, Ankita Sahu, Satyartha Prakash, Meghana Arvind, Rajat Ujjainiya, Tarani Mathur, Shilpa Ray, Ayushi Narayan, Harleen Kaur, Vishu Gupta, Mohit Kumar Divakar, Mamta Rathore, Prateek Singh, Vikas M Hiremath, Saraswati Awasthi, Kanika Singh, Divya Bhalla, Anubhuti Bansal, Yogesh Kumar, Vignesh S Kumar, Shivani Chitkara, Charvy Rana, Amit Maurya, Sreeshma Raj K, Anshul Verma, Pulkit Hasmukhbhai Leuva, Pratik Pathade, Safeer Khan, Anshul Bhardwaj, Bharti, Shail Kumari, Bharti sharma, Ruchi, Shubham Kumar, Tanmay pawaskar, Sumant Kumar, Mohit, Shyam Singh Bisht, Deeksha Yadav, Prajwali Sawant, Rohan Bhardwaj, Shailja kant Upadhyay, Rushikesh joshi, Shaivyanand Singh, Shivam Dhawan, Vanshika Srivastava, Dikkshita Baruah, Rohit Kumar, Praveena Mishra, Ankit Basnal, Md. Intyaz Ali, Pranjal Tewari, Swati, Abhishek Kumar, Nancy Rawat, Azhar uddin, Kamal singh Pindari, Deepak (**IGIB**), MK Kanakavalli, Rakhesh K V, Neha Kumari, Jukanti Akshitha, Deepshikha Esari, Mohammed Osed Annan, Mahfuj Hasan, Dibya Rana Saha Roy (**CCMB**), Ajit A Sutar, Sagar Baulia, Ameya A Pawar, Monika Sharma, Milind Kale, Ankita Namdeo, Rajesh S, Rashdajabeen Q Shaikh, Vaishnavi N Mahajan (**NCL**), Ankita Mridha, Saheli Chowdhury, Pratitusti Basu, Dipamoy Dutta, Dwaipayan Saha, Ruby Banerjee (**IICB**), Sai Adarsh Sahu, Sk Rameej Raja (**IMMT**), Abhisheik Eedara, Vishwa Priya, Navya Sahithi Pelimelli, srilekha Anumulapuri (**IICT**), Shail Singh, Lakra Promila, Swarnali Basu, Rahul Roy, Shweta Tiwari, Shikha Yadav, Amit Kumar Shahravat, Kabita Sarkar, Adrija Rakshit, Deepanshu Sindhwani, Kajal KM, Smita Pandey (**CDRI**), Sangavi Pakkyam (**CECRI**), Vipul Sharma (**CEERI**), Dikchha Singh (**CIMFR**), Parimala karupannan (**CLRI**), Ravi Raj, Ankita Kumari (**IHBT**), Nancy Sharma, Sahaurti Sharma, Sakshi Nagial (**IIIM,Jammu**), Mir Shahid Maqbool, Kaneez Fatima (**IIIM,Srinagar**), Suchismita Benjwal, Pramod Chauhan (**IIP**), Neha Mehrotra (**IITR**), Parvez Ahmad, Priyadarshan Kinatukara, Bhupender Singh, Shiva Sundharam S, Pranavathiyani G, Kuldeep Singh, Rakesh Kumar, Pradip Sen, Siddhakam Palmal, Ritu Jatav, Lalit Kumar, Pravin Kumar, Priyanshu Singh Raikwar, Simran Gambhir (**IMTECH**), Madan Mohan Pandey (**NBRI**), Manuj Kr Das, Borsha Rajkumari, Sukanya Borkakoti, Mamta Thapa, Ashique Hussain, Ishant Jyoti Nath (**NEIST**), Rahul R Menon, Sanoj Mohan, Arnold Moses (**NIIST**), Akshika (**NIScPR**), Navneet Singh Randhawa, Priyanka Singh, K Sudhakara Rao (**NML**).

## Sample aliquoting

Aastha Mishra, Ankita Sahu, Meghana Arvind, Shivani Chitkara, Deeksha Yadav, Mohit, Shyam Singh Bisht, Shail Kumari, Ankit Basnal, Ansuman Sahu, Ruchi, Tanmay Pawaskar, Nancy Rawat, Pranjal Tewari, Vignesh S Kumar, Safeer Khan, Anshul Bhardwaj (**IGIB**)

## Assays

Shantanu Sengupta, Viren Sardana, Kumardeep Chaudhary, Vamsi K. Yenamandra, Rakesh Sharma, Aastha Mishra, Vivek Rao, Shantanu Chowdhury, Beena Pillai, Sheetal Gandotra, Rajesh Pandey Ankita Sahu, Satyartha Prakash, Vignesh S Kumar, Safeer Khan, Shilpa Ray, Tarani Mathur, Rajat Ujjainiya, Mamta Rathore, Yogesh Kumar, Ankur Halder, Azhar uddin, Shivani Chitkara, Md Quasid Akhter, Bharti, Shubham Kumar, Sumant Kumar, Shail Kumari, Anshul Bhardwaj, Ruchi, Deeksha Yadav, Jitendra Kumar, Abhishek Kumar, Swati **(IGIB**), Partha Chakraborty, Dipyaman Ganguly (**IICB**). Giriraj Ratan Chandak, Swasti Raychaudhuri, Karthik Bharadwaj Tallapaka, Jukanti Akshitha, Dimple Lavanuru, Sumati Swain (**CCMB**), Mahesh J Kulkarni, Chiranjit Chowdhury, Rajesh S, Ajit A Sutar, Ameya A Pawar, Milind Kale (**NCL**), Dipyaman Ganguly, Partha Chakraborty, Saikat Majumder, Shilpak Chatterjee, Jahangir Alam (**IICB**), Umakanta Subudhi, Sai Adarsh Sahu, Sk Rameej Raja (**IMMT**) Ramakrishna Sistla, Sai Balaji Andugulapati (**IICT**), Amit Lahiri, Mrigank Srivastava, Shail Singh, Lakra Promila, Swarnali Basu, Shweta Tiwari, Shikha Yadav, Kabita Sarkar, Adrija Rakshit, Deepanshu Sindhwani (**CDRI**)

## Genomic DNA Isolation

Aastha Mishra, Ankita Sahu, Shubham Kumar, Shyam Singh Bisht, Safeer Khan, Bharti, Ruchi, Tanmay Pawaskar, Rohit Kumar, Nancy Rawat, Swati, Pranjal Tewari (**IGIB**).

## Data management and curation

Kumardeep Chaudhary, Viren Sardana, Shantanu Sengupta, Ajay Pratap Singh, Ankita Sahu, Meghana Arvind, Satyartha Prakash, Vignesh S Kumar, Pulkit Hasmukhbhai Leuva, Anshul Verma, Sreeshma Raj K, Pratik Pathade, Tarani Mathur, Kalyani Verma, Yogesh Kumar, Md. Intyaz Ali, Sudhir Rohilla (**IGIB**)

## Manuscript writing, review and editing

Shantanu Sengupta, Viren Sardana, Kumardeep Chaudhary, Vamsi K. Yenamandra, Satyartha Prakash, Vignesh S Kumar, Anshul Verma, Sreeshma Raj K, Pulkit Hasmukhbhai Leuva (**IGIB**),Giriraj Ratan Chandak, Swasti Raychaudhuri, Karthik Bharadwaj Tallapaka (**CCMB**), Mahesh J Kulkarni (**NCL**), Partha Chakraborty, Dipyaman Ganguly (**IICB**), Umakanta Subudhi (**IMMT**).

## Data Availability

Anonymized data for public use may be made available after three years from completion of baseline phase of study or as per advisory from Monitoring Committee of the project, if any revisions thereof.

## List of Abbreviations

AC: Abdominal Circumferences
ADA: American Diabetes Association
AI: Artificial Intelligence
API: Application Programming Interface
ATS: American Thoracic Society
BCA: Body Composition Analysis
BMI: Body Mass Index
CAP: Continuous Attenuation Parameter
CAP: College of American Pathologists
CC: Chest Circumferences
CLI: Command-Line Interfaces
CRF: Cryomed Controlled-Rate Freezer
CSIR: Council of Scientific and Industrial Research
CSS: Cascading Style Sheets
CTRI: Clinical Trials Registry-India
CVD: Cardiovascular Diseases
ECG: Echocardiogram
EDTA: Ethylenediamine Tetraacetic Acid
ERS: European Respiratory Society
FBS: Fasting Blood Sugar
FEV1: Forced Expiratory Volume
FRS: Framingham Risk Score
FVC: Forced Vital Capacity
GSA: Global Screening Array
GUI: Graphical User Interface
HbA1c: Glycated Hemoglobin
HC: Hip Circumferences
HRV: Heart Rate Variability
ICDS: Integrated Child Development Services
ICP-MS: Inductively Coupled Plasma Mass Spectrometer
IHEC: Institutional Human Ethics Committee
LMICs: Low and Middle-Income Countries
LSM: Liver Stiffness Measurement
ML: Machine Learning
MRM: Multiple Reaction Monitoring
NABL: National Accreditation Board for Testing and Calibration Laboratories
NAFLD: Nonalcoholic Fatty Liver Disease
NASH: Nonalcoholic Steato-Hepatitis
NCDs: Non-Communicable Diseases
OAD: Obstructive Airway Disease
PI-CheCK: Phenome India-CSIR Health Cohort Knowledgebase
qPCR: Quantitative Polymerase Chain Reaction
RBAC: Role-Based Access Control
RDBMS: Relational Database Management System
REDCap: Research Electronic Data Capture
RPM: Revolutions Per Minute
SBS: Sequence By Synthesis
SFTP: Secure File Transfer Protocol
SQL: Structured Query Language
SST: Serum-Separating Tube
TB: Terabytes
TE: Transient Elastography
TEWL: Transepidermal Water Loss
UT: Union Territories
VPN: Virtual Private Network
WC: Waist Circumferences

---

**Supplementary Table S1:**
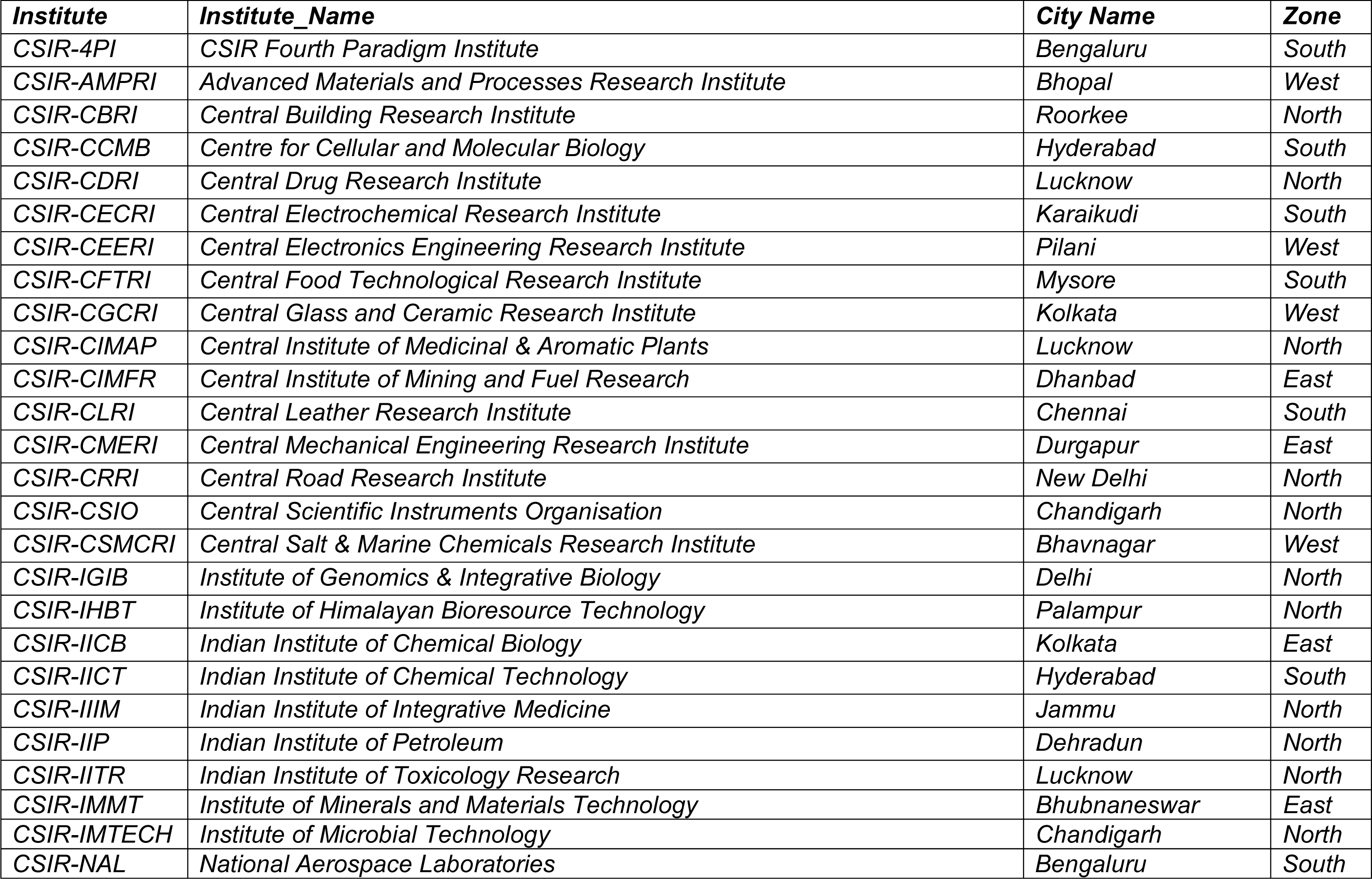
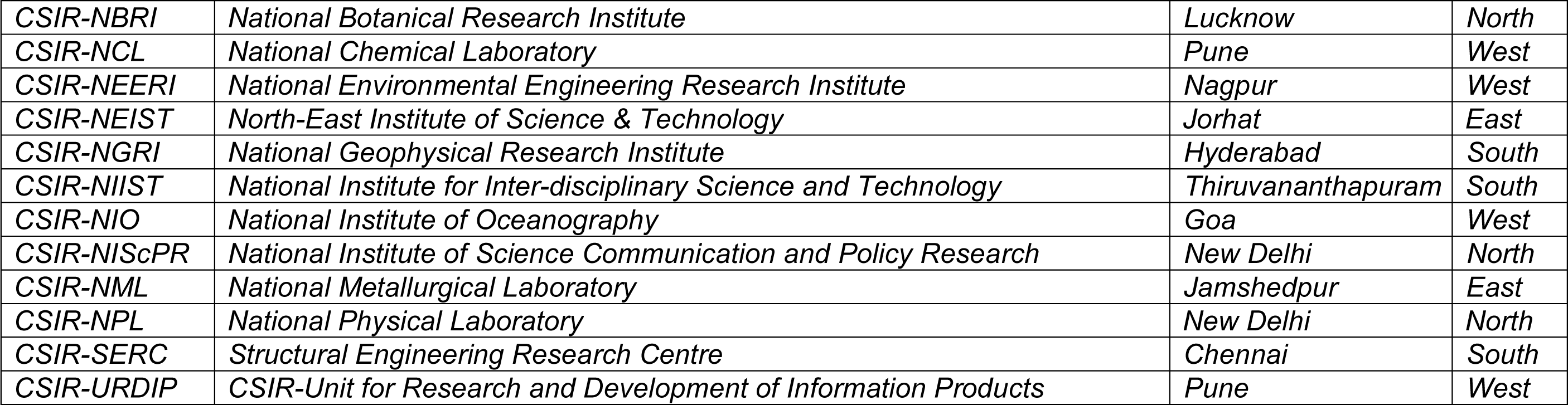
List of CSIR Institutes serving as sample collection centers stratified by zones in PI-CHeCK study.

| **Institute** | **Institute_Name** | **City Name** | **Zone** |
| --- | --- | --- | --- |
| CSIR-4PI | CSIR Fourth Paradigm Institute | Bengaluru | South |
| CSIR-AMPRI | Advanced Materials and Processes Research Institute | Bhopal | West |
| CSIR-CBRI | Central Building Research Institute | Roorkee | North |
| CSIR-CCMB | Centre for Cellular and Molecular Biology | Hyderabad | South |
| CSIR-CDRI | Central Drug Research Institute | Lucknow | North |
| CSIR-CECRI | Central Electrochemical Research Institute | Karaikudi | South |
| CSIR-CEERI | Central Electronics Engineering Research Institute | Pilani | West |
| CSIR-CFTRI | Central Food Technological Research Institute | Mysore | South |
| CSIR-CGCRI | Central Glass and Ceramic Research Institute | Kolkata | West |
| CSIR-CIMAP | Central Institute of Medicinal & Aromatic Plants | Lucknow | North |
| CSIR-CIMFR | Central Institute of Mining and Fuel Research | Dhanbad | East |
| CSIR-CLRI | Central Leather Research Institute | Chennai | South |
| CSIR-CMERI | Central Mechanical Engineering Research Institute | Durgapur | East |
| CSIR-CRRI | Central Road Research Institute | New Delhi | North |
| CSIR-CSIO | Central Scientific Instruments Organisation | Chandigarh | North |
| CSIR-CSMCRI | Central Salt & Marine Chemicals Research Institute | Bhavnagar | West |
| CSIR-IGIB | Institute of Genomics & Integrative Biology | Delhi | North |
| CSIR-IHBT | Institute of Himalayan Bioresource Technology | Palampur | North |
| CSIR-IICB | Indian Institute of Chemical Biology | Kolkata | East |
| CSIR-IICT | Indian Institute of Chemical Technology | Hyderabad | South |
| CSIR-IIIM | Indian Institute of Integrative Medicine | Jammu | North |
| CSIR-IIP | Indian Institute of Petroleum | Dehradun | North |
| CSIR-IITR | Indian Institute of Toxicology Research | Lucknow | North |
| CSIR-IMMT | Institute of Minerals and Materials Technology | Bhubaneswar | East |
| CSIR-IMTECH | Institute of Microbial Technology | Chandigarh | North |
| CSIR-NAL | National Aerospace Laboratories | Bengaluru | South |
| *CSIR-NBRI* | *National Botanical Research Institute* | *Lucknow* | *North* |
| *CSIR-NCL* | *National Chemical Laboratory* | *Pune* | *West* |
| *CSIR-NEERI* | *National Environmental Engineering Research Institute* | *Nagpur* | *West* |
| *CSIR-NEIST* | *North-East Institute of Science & Technology* | *Jorhat* | *East* |
| *CSIR-NGRI* | *National Geophysical Research Institute* | *Hyderabad* | *South* |
| *CSIR-NIIST* | *National Institute for Inter-disciplinary Science and Technology* | *Thiruvananthapuram* | *South* |
| *CSIR-NIO* | *National Institute of Oceanography* | *Goa* | *West* |
| *CSIR-NIScPR* | *National Institute of Science Communication and Policy Research* | *New Delhi* | *North* |
| *CSIR-NML* | *National Metallurgical Laboratory* | *Jamshedpur* | *East* |
| *CSIR-NPL* | *National Physical Laboratory* | *New Delhi* | *North* |
| *CSIR-SERC* | *Structural Engineering Research Centre* | *Chennai* | *South* |
| *CSIR-URDIP* | *CSIR-Unit for Research and Development of Information Products* | *Pune* | *West* |

**Supplementary Table S2:**
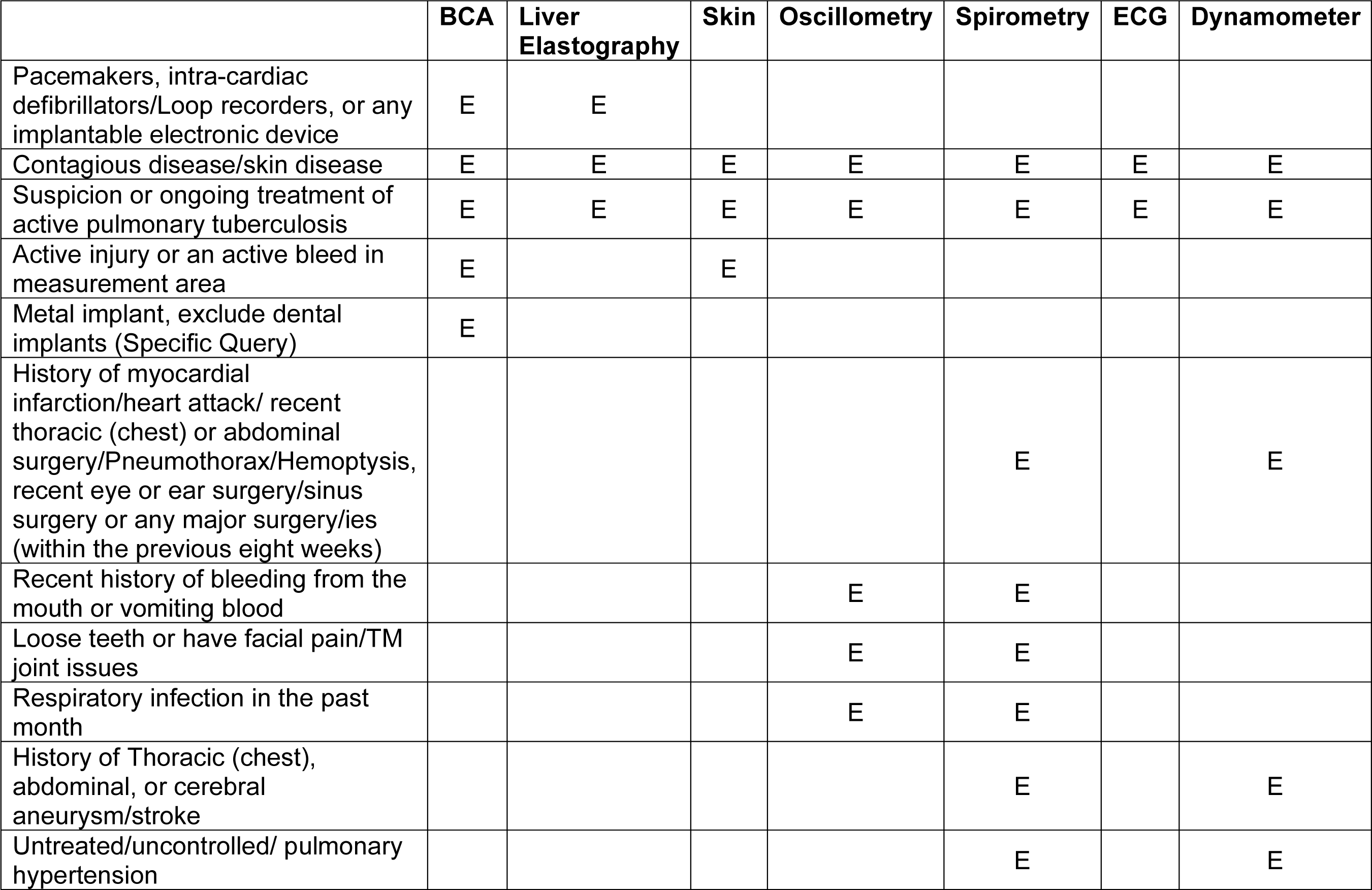
Details of exclusion (E) criteria for different scanning modalities.

|  | **BCA** | **Liver Elastography** | **Skin** | **Oscillometry** | **Spirometry** | **ECG** | **Dynamometer** |
| --- | --- | --- | --- | --- | --- | --- | --- |
| Pacemakers, intra-cardiac defibrillators/Loop recorders, or any implantable electronic device | E | E |  |  |  |  |  |
| Contagious disease/skin disease | E | E | E | E | E | E | E |
| Suspicion or ongoing treatment of active pulmonary tuberculosis | E | E | E | E | E | E | E |
| Active injury or an active bleed in measurement area | E |  | E |  |  |  |  |
| Metal implant, exclude dental implants (Specific Query) | E |  |  |  |  |  |  |
| History of myocardial infarction/heart attack/ recent thoracic (chest) or abdominal surgery/Pneumothorax/Hemoptysis, recent eye or ear surgery/sinus surgery or any major surgery/ies (within the previous eight weeks) |  |  |  |  | E |  | E |
| Recent history of bleeding from the mouth or vomiting blood |  |  |  | E | E |  |  |
| Loose teeth or have facial pain/TM joint issues |  |  |  | E | E |  |  |
| Respiratory infection in the past month |  |  |  | E | E |  |  |
| History of Thoracic (chest), abdominal, or cerebral aneurysm/stroke |  |  |  |  | E |  | E |
| Untreated/uncontrolled/ pulmonary hypertension |  |  |  |  | E |  | E |

**Supplementary Table S3:**
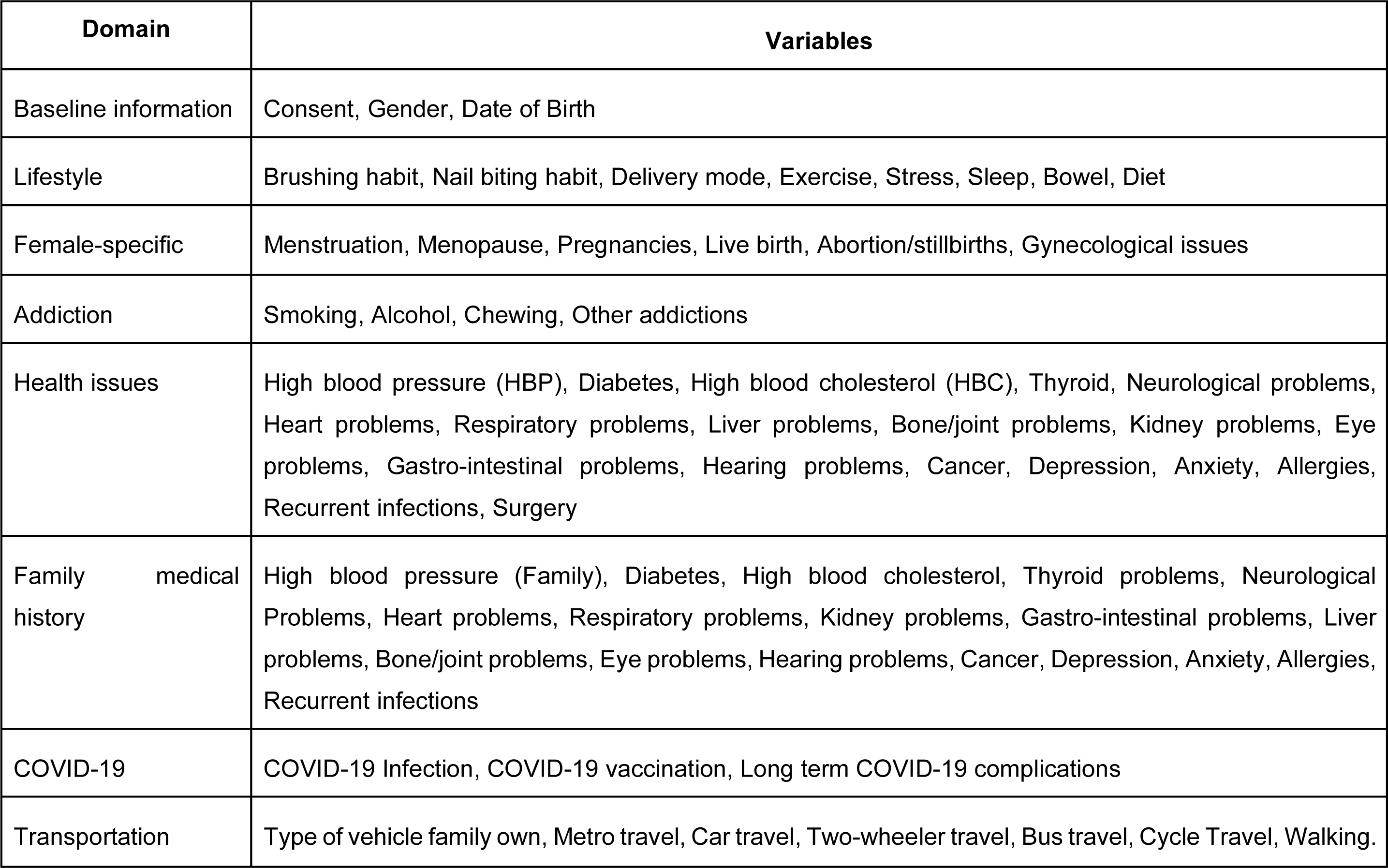
Description of self-reported Main questionnaire.

**Supplementary Table S4:** Description of the self-reported Gut questionnaire.

| Domain | Variables |
| --- | --- |
| Food item consumption | Rice, Wheat (Paratha, chapati, roti, etc), Other cereals (Bajra, jowar, ragi etc), Bread(Yeast), Pulses (Arhar, tur, moong, urd daal etc), Sprouts, Eggs, Poultry (Chicken), Meat (Mutton, pork etc), Fish and other seafoods, Curd, Paneer, Ghee, Butter, Ice cream, Cake and pastries, Chips, namkeen, Pickles, Biscuits, Chocolate, Carbonated beverages (Coke, sprite etc), Fruit juice and shakes, Tubers and starchy vegetables (Potato, corn etc), Green leafy vegetables, Fruits (Banana, apple etc), Dry fruits (Resins, almonds, dates etc), Fermented foods (Idli, dosa, dhokla etc), Fermented drinks, Vitamin supplements, Protein supplements, Probiotics |
| Alcohol | Beer consumption within 2 days, Alcohol consumption within two days |
| Travel & eating | Types of green leafy vegetables consumed last week, Travel out of city or country in the last 15 days. |
| Antibiotic consumption | Last antibiotic consumption, Antibiotics consumption within two weeks |
| Hospitalization | Surgery in last one month, Hospitalized in last one month |
| Stool type | Bristol stool type |

**Supplementary Table S5:** Summary of different sampling/scanning stations.

| **Station (Modality)** | **Machine** | **Information** |
| --- | --- | --- |
| Anthropometry | Dynamometer- Baseline | Muscle strength |
| BCA | BIA Analyzer-3 Frequency<br>ACCUNIQ BC380 | Body Composition |
| Phlebotomy | Details in Supplementary Table S6 | Blood parameters |
| Transient Elastography | Fibroscan Mini 430Plus | LSM and CAP score |
| ECG | Allengers Pisces 1012 | Heart condition |
| Spirometry | NDD Easy One Air | Lung Function and Volume |
| Oscillometry | Tremoflo C100, Resmon Pro V3 | Small Airway Assessment |
| Skin | Courage Khazaka | Skin health |

**Supplementary Table S6:** Details of the phlebotomy parameters, including biochemistry assays.

| **Diagnostic Lab-Outsourced** |  |  |  |  |  |
| --- | --- | --- | --- | --- | --- |
| **S. No.** | **Units** | **Parameter** | **Sample Type** | **Platform** | **Sample Size** |
| 1 | 10<sup>3</sup>/uL | Absolute Basophil Count | WB | DHBC | All |
| 2 | 10<sup>3</sup>/uL | Absolute Eosinophil Count (AEC) | WB | DHBC | All |
| 3 | 10<sup>3</sup>/uL | Absolute Lymphocyte Count (ALC) | WB | DHBC | All |
| 4 | 10<sup>3</sup>/uL | Absolute Monocyte Count | WB | DHBC | All |
| 5 | 10<sup>3</sup>/uL | Absolute Neutrophil Count (ANC) | WB | DHBC | All |
| 6 | U/L | Alanine Aminotransferase (ALT/SGPT) | Serum | AUBC | All |
| 7 | Ratio | Albumin/Globulin Ratio | Serum | AUBC | All |
| 8 | U/L | Alkaline Phosphatase (ALP) | Serum | AUBC | All |
| 9 | mg/dL | Apolipoprotein A1 (APO-A1) | Serum | AUBC | All |
| 10 | mg/dL | Apolipoprotein B (APO-B) | Serum | AUBC | All |
| 11 | U/L | Aspartate Aminotransferase (AST/SGOT) | Serum | AUBC | All |
| 12 | mg/dL | Average Estimated Glucose - plasma | Calculated | DHBC | All |
| 13 | % | Basophils | Serum | DHBC | All |
| 14 | mg/dL | Blood Urea | Serum | AUBC | All |
| 15 | mg/dL | Blood Urea Nitrogen (BUN) | Serum | AUBC | All |
| 16 | Ratio | Bun/Creatinine Ratio | Calculated | AUBC | All |
| 17 | % | Eosinophils | WB | DHBC | All |
| 18 | mm/1st hour | Erythrocyte Sedimentation Rate (ESR) | WB | ALIFAX TEST-1 | All |
| 19 | ng/mL | Ferritin | Serum | ATELLICA IM (SIEMENS) | All |
| 20 | U/L | Gamma Glutamyl Transferase (GGT) | Serum | AUBC | All |
| 21 | mL/min/1.73m<sup>2</sup> | estimated glomerular filtration rate (eGFR) | Calculated | AUBC | All |
| 22 | mg/dL | Glucose, Fasting | Fluoride | AUBC | All |
| 23 | Ratio | Green and king index | WB | DHBC | All |
| 24 | g/dL | Haemoglobin (HB) | WB | DHBC | All |
| 25 | % | HbA1c (Glycosylated Hemoglobin) | WB | BDB | All |
| 26 | Ratio | High-density lipoprotein (HDL)/low-density lipoprotein (LDL) Cholesterol Ratio | Serum | AUBC | All |
| 27 | % | Hematocrit (PCV) | WB | DHBC | All |
| 28 | Ratio | LDL/HDL Cholesterol Ratio | Serum | AUBC | All |
| 29 | % | Lymphocytes | WB | DHBC | All |
| 30 | pg | Mean Corp Hb (MCH) | WB | DHBC | All |
| 31 | g/dL | Mean Corp Hb Conc (MCHC) | WB | DHBC | All |
| 32 | fL | Mean Corp Volume (MCV) | WB | DHBC | All |
| 33 | Ratio | Mentzer Index | WB | DHBC | All |
| 34 | % | Monocytes | WB | DHBC | All |
| 35 | fL | MPV | WB | DHBC | All |
| 36 | % | Neutrophils | WB | DHBC | All |
| 37 | mg/dL | Non-HDL Cholesterol | Serum | AUBC | All |
| 38 | 10<sup>3</sup>/uL | Platelet Count (PLT) | WB | DHBC | All |
| 39 | % | Red Cell Distribution Width - Coefficient of Variation (RDW-CV)– | WB | DHBC | All |
| 40 | fL | Red Cell Distribution Width-Standard Deviation (RDW - SD) | WB | DHBC | All |
| 41 | Ratio | Red Cell Distribution Width Index (RDWI) | WB | DHBC | All |
| 42 | 10<sup>6</sup>/uL | Red Blood Cell Count (RBC) | WB | DHBC | All |
| 43 | g/dL | Serum Albumin | Serum | AUBC | All |
| 44 | mg/dL | Serum Bilirubin, (Direct) | Serum | AUBC | All |
| 45 | mg/dL | Serum Bilirubin, (Indirect) | Serum | AUBC | All |
| 46 | mg/dL | Serum Bilirubin, (Total) | Serum | AUBC | All |
| 47 | mg/dL | Serum Calcium | Serum | AUBC | All |
| 48 | mmol/L | Serum Chloride | Serum | AUBC | All |
| 49 | mg/dL | Serum Creatinine | Serum | AUBC | All |
| 50 | gm/dL | Serum Globulin | Serum | AUBC | All |
| 51 | mg/dL | Serum HDL Cholesterol | Serum | AUBC | All |
| 52 | ug/dL | Serum Iron | Serum | AUBC | All |
| 53 | mg/dL | Serum LDL Cholesterol | Serum | AUBC | All |
| 54 | mg/dL | Serum Phosphorus | Serum | AUBC | All |
| 55 | mmol/L | Serum Potassium | Serum | AUBC | All |
| 56 | mmol/L | Serum Sodium | Serum | AUBC | All |
| 57 | µg/dL | Serum Total Iron Binding Capacity (TIBC) | Serum | AUBC | All |
| 58 | g/dL | Serum Total Protein | Serum | AUBC | All |
| 59 | mg/dL | Serum Triglycerides | Serum | AUBC | All |
| 60 | mg/dL | Serum Uric Acid | Serum | AUBC | All |
| 61 | mg/dL | Serum very low-density lipoprotein (VLDL) Cholesterol | Serum | AUBC | All |
| 62 | Ratio | SGOT/SGPT Ratio | Calculated | AUBC | All |
| 63 | uIU/mL | Thyroid Stimulating Hormone (TSH)-<br>Ultrasensitive | Serum | AIS | All |
| 64 | Ratio | Total CHOL/HDL Cholesterol Ratio | Calculated | AUBC | All |
| 65 | mg/dL | Total Cholesterol | Serum | AUBC | All |
| 66 | 10<sup>3</sup>/uL | Total Leucocyte Count (TLC) | WB | DHBC | All |
| 67 | % | Transferrin Saturation % | Serum | AUBC | All |
| 68 | pg/mL | Unsaturated iron binding capacity (UIBC) | Serum | AUBC | All |
| 69 | Ratio | Urea/Creatinine Ratio | Calculated | AUBC | All |
| 70 | pg/mL | Vitamin B12 | Serum | AIS | All |
| 71 | ng/mL | Vitamin D (25-OH Vitamin D) | Serum | AIS | All |

WB: Whole Blood; DHBC: DxH-900 Beckman Coulter; AUBC: AU5800 Beckman Coulter; BDB: BIORAD D-100 (BIORAD); AIS: ATELLICA IM (SIEMENS)

**Supplementary Table S7:**
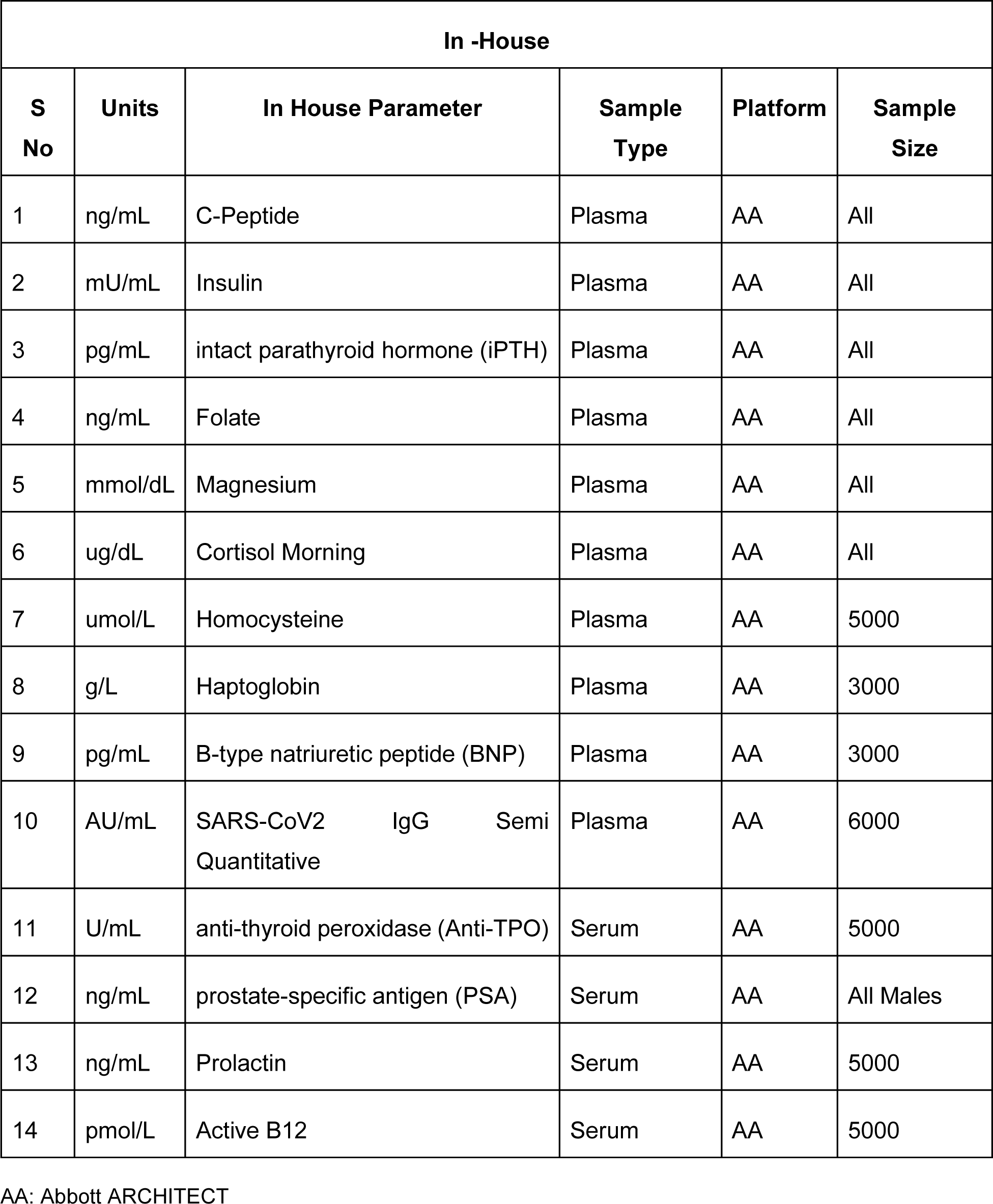
Details of the phlebotomy parameters, including biochemistry assays (In -House).

